# Global Distribution and Characteristics of Research Facilities Participating in Phase III Oncology Trials

**DOI:** 10.64898/2026.02.08.26345855

**Authors:** Felippe Lazar Neto, Rafael Trindade Santos Costa, Amanda Ferreira Villarino, Flavio Lazar, João Wilson da Rocha, Fabio Ynoe de Moraes, Jose Mauricio Mota

## Abstract

**Background:** Research infra-structure is essential for conducting phase III cancer clinical trials as its lack precludes trial availability and its maturity may influence the portfolio of available options. Several studies have highlighted global disparities, but none has ever mapped the worldwide network of research facilities conducting such trials.

**Methods:** We extracted all research sites within recruiting phase III cancer interventional trials from the ClinicalTrials.gov database on July 23, 2024. Address components were combined and queried through the Google Maps API for standardized identification. Matched pairs were subsequently screened for inconsistencies, and Google Maps entries within 1,000 meters were grouped if they represent the same facility. We compared research facilities’ number, density (per 1M inhabitants), size, and portfolio of available trials across World Development Index regions, and modelled the number of available trials and research facilities per country with log-log linear models (elasticity coefficient [β] estimand).

**Findings:** Of 77,625 listed sites from 1,287 trials, 65,736 (84.7%) were mapped to 6,634 unique research facilities across 84 countries. We found a strong correlation between the number of research facilities and trials by country (R^2^=0.86, p-value<0.001, β=0.95, 95%CI 0.87-1.04). The United States and China had the largest number of facilities (2,626, 39.6%) and available trials (624, 48.5%) respectively. All the 100 largest research facilities in size were from high-income countries or China, and 16 of the top 20 countries by density were located in Europe and Central Asia. Latin America and Caribbean, South Asia Region, and Sub-Saharan Africa had the highest proportion of facilities running only multiregional (>93%, p-value<0.001), industry-sponsored (>90%, p-value<0.001), and systemic therapy trials (>80%, p-value<0.001).

**Interpretation:** Phase III cancer trials are directly limited and responsive to physical institutional growth. Outside high-income countries, research capacity remains substantially constrained, largely focused on systemic therapies and frequently reliant on industry-sponsored, multiregional trials.

**Funding:** None

**RESEARCH IN CONTEXT:** *Evidence before this study:* We searched the Pubmed/EMBASE database for studies investigating cancer clinical trials and research facilities distribution worldwide using the search terms “trials OR clinical trials”, “cancer”, “distribution OR barriers OR access OR inequalit*” between inception to November 1st, 2025. Several studies have consistently shown a higher concentration and availability of cancer clinical trials in high-income countries (HIC) compared to lower-income regions. In addition, cancer clinical trials in lower middle-income countries (LMIC) are predominantly phase III, industry-sponsored, and led by HIC. However, most of this evidence is derived from country-level aggregate data, which collapse heterogeneous research infrastructures within countries into single summary measures and therefore lack the resolution needed to describe, and ultimately explain the underlying drivers of these disparities. Only a small number of studies have examined cancer trial distribution at finer geographic scales, and these have generally been limited to city or county-level analyses or to single-country settings. Therefore, there remains no comprehensive mapping of the research infrastructure responsible for conducting cancer clinical trials, even though this infrastructure is a key determinant of national trial availability and study profiles.

*Added value of this study:* This is the first study to map and profile research facilities conducting phase III cancer clinical trials worldwide. We show that the availability of phase III cancer clinical trials is linearly correlated with the absolute number of research facilities on a log-log scale, such that a 1% increase in research facilities is associated with a 0.95% (95%CI, 0.87–1.04) increase in phase III trial availability within countries. The United States had the largest share of research facilities (N=2,626, 39%) while China had the highest number of available phase III clinical trials (N=624), driven by a high number of single-center studies (N=398). Of the largest 100 research facilities in size (median [IQR] number of phase III cancer trials 62 [57-73.25]), all were from HIC or China. Of the top 20 countries by density (per million people), 80% were located in Europe and Central Asia (ECA). In Latin America and Caribbean (LAC), South Asia Region (SAR) and Sub-Saharan Africa (SSA), research facilities are found in fewer numbers and smaller sizes, and are predominantly running only multiregional (>93%), industry-sponsored (>90%), and systemic therapy trials (>80%).

*Implications of all the available evidence:* Global disparities in cancer clinical trials are perpetuated by constrained research infrastructure in less developed regions, reflected not only in the limited number of research facilities but also in their profiles, which often have little experience beyond industry-sponsored trials, as we have shown. While investments in research facilities are crucial and are associated with increased trial availability, particularly during the early stages of infrastructure development, they are not sufficient on their own. Complementary strategies are needed, including financial incentives to support locally designed trials (i.e., investigator-initiated grants) and sustained investment in human resources to design and conduct them. Aligning infrastructure expansion with workforce development is essential to improve both the quantity and the profile of available cancer clinical trials, enhance local leadership, and ensure that research is relevant and generalisable to diverse populations.

## INTRODUCTION

Cancer is the first and second leading cause of death in 57 and 70 countries^1^, respectively, and its incidence is projected to rise over the next decades due to improved diagnosis and shifting age demographics^2^. This rise will disproportionately affect developing regions where there is more room for improvement in health access and a larger shift in the demographic pyramid is expected^2^. In contrast, cancer mortality has consistently decreased over the past 20 years in developed countries^3^. This can be explained by multiple factors, one being the introduction of more effective cancer treatments. From 2003 to 2021, the United States (US) Food and Drug Administration (FDA) approved 124 cancer drugs comprising 374 indications, with averaged improved survival hazards of 27% among 234 randomized clinical trials with survival data available^4^.

Clinical trials are the gold standard experimental design to test new therapies that will ultimately be approved for routine clinical use. Despite the overall positive beliefs from the patients’ perspective^5,6^, the proportion of patients enrolled in clinical trials remains very low (≤ 10%), particularly outside academic centers^7^. In addition, recruited patients often do not resemble regional populations, consistently under-representing ethnic minorities^8–11^, women^12^, and patients from socially deprived areas^13^, despite some modest recent improvement^8^. Common barriers for recruitment include lack of knowledge^5,14,15^, travel distance^6,16^, fear of treatment side effects^6,15^, financial constraints^6,17^, restricted selection criteria^18^, and lack of clinical trial options^19–21^. In fact, the large majority of patients won’t be enrolled because no trials are available near them.

Cancer clinical trials availability disparities can happen both at the national^19,20,22^ and global level^23–25^. For example, in the US, where a large number of trials are often available, 70% of the counties had no active trials in 2022, encompassing 74% of the US land^20^. At the global level, 60 out of 175 countries had no trials available, and only 56 countries had trials registered every year^25^. Additionally, several studies have systematically shown large differences between high- and lower-income countries’ clinical trials availability that go beyond the absolute number of studies to describe a higher proportion of industry-funded in lower-income countries^23,24,26,27^. A systematic review of common barriers to conducting clinical trials in developing countries highlighted many of the possible reasons, including financial limitations, ethical and regulatory obstacles, and lack of research infrastructure^28^.

Research infra-structure is fundamental for conducting clinical trials as the lack of research facilities precludes trial availability. Previous studies have investigated cancer trials density at the global^23–25^, and country^19,20,22^ level, but none has ever tried to map the worldwide network of research facilities conducting cancer clinical trials. This information is relevant because it can provide global insights into the quantity and profile of research infrastructure in each country which might be determinant for the portfolio of available trials. Leveraging the comprehensiveness of the ClinicalTrials.Gov database and the extensive global coverage of Google Maps^29^, we aimed to map, describe and analyze the research infrastructure conducting phase III cancer clinical trials worldwide.

## METHODS

### Study Design and Database Search Strategy

We performed a cross-sectional analysis of all clinical trials with “cancer” as “condition” and “overall status” as “recruiting” from the ClinicalTrials.Gov database using their application programming interface (API) (https://clinicaltrials.gov/api/v2/studies) on 23th, July, 2024. For every trial, we extracted study identification number (NCT), title, eligibility criteria, conditions, design type, design phase, primary purpose, sponsor lead name, sponsor type and all their listed locations, including facility name, formatted address, city, state, zip code, country and recruitment status. This preliminary database was further filtered to include only trials listed as design “phase III”, design type “interventional”, and primary purpose “treatment” or “supportive care”. Trial information was manually reviewed, and those with cancer as not the main research focus (e.g., “parathyroidism”) were excluded. We chose the ClinicalTrials.Gov database given it is the most comprehensive database on clinical trials with available granular data on recruiting locations (e.g. name, city) to potentially identify them.

### Research Facilities Identification

For every listed location, we created a unique identifier using the combination of its facility name, city, state, zip code and country, after text sanitization. After duplicate identifiers were removed, unique locations were queried through the Google Maps API (https://maps.googleapis.com/maps/api/place/findplacefromtext/json), using the full address string formatted as ‘facility name, city, state, zip code, country’. For each matched candidate, the API returned candidate places metadata including the Google place identification, the formatted address, country, and geocoordinates (latitude and longitude). Every pair of unique locations and Google places were manually reviewed. Locations where the returned Google places did not correspond to the address location, were determined as unidentified. Facility locations with generic names (e.g., “Research Site”, “Site Number: 000,” “Company Study Site”) were initially classified as “unidentified” and were not queried through the Google Maps API.

To address the potential bias of a single location having two or more Google places addresses (e.g. Department of Oncology, University; and Department of Obstetrics; University), we performed a pair-wise evaluation followed by grouping within the same facilities. First, we retrieved all candidate Google addresses within 1,000 meters of Haversine distance using their geocoordinates and separated them into pairs (A and B, B and C, and A and C, if A, B, and C are within 1,000 meters of distance). Second, two authors (AFV and RTC) manually reviewed each pair of Google addresses and determined if they corresponded to the same research facility. However, because of the highly specific geographic knowledge needed for this step, and language barriers for some institution names (e.g. non-latin alphabet), we used a large language model (LLM) as an additional reviewer. We chose the Gemini Pro 3.0 because of its performance, thinking capabilities, and its Google search tool calling which allows search of the Google knowledge database to provide answers^30^. Using prompt engineering techniques, we iteratively optimized a prompt against a random sample of 80 location pairs from Brazil, where the authors had sufficient knowledge to provide a gold standard (**Suppl. Table 1)**. We used a temperature of zero and a “high” thinking level. The model achieved accuracy of 98.8% (N=79/80 correct answers). We evaluated the output stability by repeatedly querying it with identical prompts (N=10), achieving a mean accuracy of 96.86% (SD=1.1%). Finally, we performed the research facilities clustering according to the classified pairs from Gemini Pro 3.0. We chose to use the Gemini Pro 3.0 evaluation rather than the authors’ manual review due to the model’s high accuracy and to minimize potential bias from human evaluation, particularly given the geographic and linguistic limitations inherent in manual review across diverse international institutions. The prompt and complete methodology used is described in the **Suppl. Appendix**.

**TABLE 1.** Characteristics of 6,634 research facilities participating in Phase 3 clinical trials. Continuous variables are presented as median [interquartile range]; categorical variables as N (%).

| Characteristics | Overall |
| --- | --- |
| **N** | 6634 |
| **Listed Trials, Median (IQR)** | 4.00 [1.00, 12.00] |
| **Listed Trials Category, N (%)** |  |
| 1-5 Trials | 3874 (58.4) |
| 6-10 Trials | 908 (13.7) |
| 11-20 Trials | 875 (13.2) |
| 21-50 Trials | 849 (12.8) |
| 51+ Trials | 128 (1.9) |
| **Any Single Center Trial, N (%)** | 211 (3.2) |
| **Any Radiotherapy Trial, N (%)** | 2249 (33.9) |
| **Any Surgical Trial, N (%)** | 535 (8.1) |
| **Only Multicenters Trials, N (%)** | 6423 (96.8) |
| **Only Multinational Trials, N (%)** | 3367 (50.8) |
| **Only Multiregional Trials, N (%)** | 2802 (42.2) |
| **Only Industry-Sponsored Trials, N (%)** | 2572 (38.8) |
| **Only Systemic-Therapy Trials, N (%)** | 3468 (52.3) |
| **Country, N (%)** |  |
| United States | 2626 (39.6) |
| China | 464 (7.0) |
| Germany | 383 (5.8) |
| France | 273 (4.1) |
| Japan | 263 (4.0) |
| United Kingdom | 230 (3.5) |
| Italy | 190 (2.9) |
| Brazil | 157 (2.4) |
| Australia | 152 (2.3) |
| Spain | 134 (2.0) |
| Poland | 124 (1.9) |
| Canada | 98 (1.5) |
| Russian Federation | 97 (1.5) |
| India | 96 (1.4) |
| Turkey | 89 (1.3) |
| Others (69 countries) | 1173 (17.6) |
| **WDI Regions Involved, N (%)** |  |
| North America | 2724 (41.1) |
| Europe & Central Asia | 2192 (33.0) |
| East Asia & Pacific | 1095 (16.5) |
| Latin America & Caribbean | 434 (6.5) |
| South Asia | 96 (1.4) |
| Middle East, North Africa, Afghanistan & Pakistan | 57 (0.9) |
| Sub-Saharan Africa | 36 (0.5) |
| **WDI Income Level, N (%)** |  |
| High income | 5343 (81.0) |
| Upper middle income | 1103 (16.7) |
| Lower middle income | 150 (2.3) |
| Low income | 2 (0.0) |

### Cancer Clinical Trials and Country Metadata

For every trial, we reviewed and manually annotated the following variables: cancer diagnosis, treatment modality (surgery, radiotherapy, systemic therapy, symptom management or others) and sponsor type (cooperative groups, government agencies, research facilities and industry). When two or more cancer diagnoses were targeted in the same trial, we classified it as a “multicancer” trial. We queried countries’ metadata from the World Development Indicators (WDI), a curated database from the World Bank. For each country, we extracted the most updated available data of the following variables: WDI region (East Asia & Pacific [EAP], Europe & Central Asia [ECA], Latin America & Caribbean [LAC], Middle East & North Africa, Afghanistan & Pakistan [MENA], North America [NAM], South Asia [SAR], Sub-Saharan Africa [SSA]), income level (high-income [HIC], upper-middle income [UMIC], lower-middle income countries [LMIC], and low-income [LIC]), gross domestic product (GDP), GDP-per capita, physicians density (per 1000 inhabitants), research expenditure value (% GDP), health expenditure (% GDP) and total population.

All data sources were combined into a single consolidated database of cancer clinical trials, locations (identified and unidentified) and their respective metadata. Clinical trials were then classified into single-center (or multicenter), single-country (or multinational) and single-region (or multiregional). We calculated the number of listed locations (unidentified and identified), number of countries, and number of regions for each trial. From the identified research facilities, we calculated the number of listed trials, and the proportion of single-centers, industry-funded, multinational and multiregional trials in each. At the country-level, we calculated the number of trials available, unique identified facilities, the facilities density (per 1M inhabitants), and the proportion of facilities with the following: any single-center trial; any radiotherapy trial, any surgery trial, only industry-funded trials; only multinational trials; and only multiregional countries. For the facilities and country level analysis, unidentified locations were excluded. In all analysis, we decided to include all listed locations irrespective of recruitment status (e.g. recruiting, not yet recruiting, completed) to ensure comprehensiveness and avoid underestimation of small or new research facilities where few trials are listed and if none are currently recruiting, it would be consequently excluded. Our ClinicalTrials.Gov search included only the overall status of active clinical trials even if some locations were not actively recruiting.

### Sensitivity Analysis

To maximize center identifications and consequently avoid potential bias with missing data, we performed a sensitivity analysis including an additional step in which we re-classified unidentified locations by their zip code. First, using the identified locations, we created a mapping of zip-codes and country to the final assigned Google places address. Second, we excluded from the mapping the zip codes where two or more Google places were matched. Finally, we applied the mapping of unique zip codes, country and Google places to the unidentified locations. Because of the potential misclassification only by the zip code, and the under-estimation of centers that are located within the same zip code (and therefore excluded from mapping), we decided to report this analysis as a sensitivity and not the main analysis.

### Statistical analysis

Numerical data were summarized using the median and interquartile range (IQR), while categorical variables were presented as absolute numbers (N) and percentages (%). We used a multilevel logistic regression model (with a random intercept for clinical trial identifier) to investigate the pattern of unidentified locations (“missingness”). Specifically, we examined whether WDI regions or sponsor status influenced the likelihood of location identification. We performed this missingness analysis in the main and sensitivity analysis data. Differences in the characteristics of clinical trials and research centers across WDI regions worldwide were evaluated using the chi-square or Fisher’s exact test for categorical variables and the Kruskal-Wallis test (non-parametric) for continuous variables.

To investigate the relationship between clinical trials availability and country research capacity, we modelled the number of trials and number of research facilities with linear regression models. We applied log-transformations to account for the right-skewed distribution of the data and visually inspected its fit. Model diagnostics included assessment of residual normality (Shapiro-Wilk test), heteroskedasticity (Breusch-Pagan test), and influential observations (Cook’s distance). Models’ goodness of fit before and after log-transformations were evaluated using the R-squared. For the resulting log-log regression model, the estimated coefficient is interpreted as the elasticity coefficient: the percentage change in the dependent variable resulting from a one percent increase in the independent variable. We also evaluated the correlation between WDI variables and research capacity (absolute number) per country using both Pearson (parametric) and Spearman-rank (non-parametric) based correlations, the latter to address possibly non-linear correlations. The level of statistical significance (alpha) for all hypothesis tests was set at 5%. All statistical analyses were conducted using R (version 4.3.3). The code is available at https://github.com/felippelazar/CancerTrialsFacilitiesGlobalDistribution/.

### Role of Funding Source

None.

## RESULTS

### Cancer Clinical Trials Characteristics

Of 19,523 recruiting cancer clinical trials, 1,281 phase III interventional trials were included, encompassing 77,625 non-unique research sites worldwide (**Figure 1**). The median (IQR) number of research sites per trial was 3 (1-56), with 44% of trials having a single site (single-centers), while 26% had 51 sites or more (**Suppl. Table 2**). China had the highest number of available trials (624, 48.5%). Nearly 30% of trials were multinational and 25% multiregional. While nearly half of trials included a HIC (593/1281, 52%) or UMIC site (586/1281, 52%), less than 5% included a LMIC and less than 0.1% a LIC. The majority of trials studied systemic treatments interventions (67%), 34% had industry sponsors, and the most common studied cancers were breast (16%), non-small cell lung cancer (10%) and colorectal (7.5%). Single-center trials (572, 44%) were predominantly found in China (398/572, 70%), followed by the United States (50/572, 8.7%) and France (19/572, 3.3%) (**Suppl. Table 2**). Among multinational trials (391, 30%), the large majority included NAM (317/391, 81%), ECA (316/391, 81%) and EAP (282/391, 72%), but only half (184/391, 47%) included LAC, and a minority of them included MENA (129/391, 33%) or the SSA (59/391, 15%) (**Suppl. Table 4)**.

**TABLE 2.** Comparison of Research Facility Characteristics by World Bank Development Indicators Region. P-values are from Kruskal-Wallis test for continuous variables and Fisher exact test for categorical variables. EAP = East Asia & Pacific; ECA = Europe & Central Asia; LAC = Latin America & Caribbean; MENA = Middle East, North Africa, Afghanistan & Pakistan; NAM = North America; SAR = South Asia; SSA = Sub-Saharan Africa.

| Characteristics | EAP | ECA | LAC | MENA | NAM | SAR | SSA | p-value |
| --- | --- | --- | --- | --- | --- | --- | --- | --- |
| **Number of Countries, N** | 15 | 41 | 12 | 11 | 2 | 1 | 5 |  |
| **Number of Research Facilities, N** | 1095 | 2192 | 434 | 57 | 2724 | 96 | 36 |  |
| **Number of Trials per Center, Median (IQR)** | 3.00 [1.00, 8.00] | 3.00 [1.00, 7.00] | 2.00 [1.00, 4.00] | 2.00 [1.00, 7.00] | 8.00 [2.00, 19.00] | 1.00 [1.00, 2.00] | 3.00 [1.00, 4.00] | <0.001 |
| **Research Facilities Density, Median (IQR)** | 3.00 [1.00, 8.00] | 3.00 [1.00, 7.00] | 2.00 [1.00, 4.00] | 2.00 [1.00, 7.00] | 8.00 [2.00, 19.00] | 1.00 [1.00, 2.00] | 3.00 [1.00, 4.00] | <0.001 |
| **Number of Trials Category, N (%)** |  |  |  |  |  |  |  | <0.001 |
| 1-5 Trials | 725 (66.2) | 1503 (68.6) | 349 (80.4) | 40 (70.2) | 1134 (41.6) | 91 (94.8) | 32 (88.9) |  |
| 6-10 Trials | 141 (12.9) | 283 (12.9) | 43 (9.9) | 5 (8.8) | 431 (15.8) | 4 (4.2) | 1 (2.8) |  |
| 11-20 Trials | 114 (10.4) | 211 (9.6) | 30 (6.9) | 5 (8.8) | 511 (18.8) | 1 (1.0) | 3 (8.3) |  |
| 21-50 Trials | 91 (8.3) | 170 (7.8) | 12 (2.8) | 2 (3.5) | 574 (21.1) | 0 (0.0) | 0 (0.0) |  |
| 51 or more Trials | 24 (2.2) | 25 (1.1) | 0 (0.0) | 5 (8.8) | 74 (2.7) | 0 (0.0) | 0 (0.0) |  |
| **Any Single Center Trial, N (%)** | 102 (9.3) | 50 (2.3) | 6 (1.4) | 11 (19.3) | 36 (1.3) | 6 (6.2) | 0 (0.0) | <0.001 |
| **Any Radiatotherapy Trial, N (%)** | 266 (24.3) | 409 (18.7) | 23 (5.3) | 11 (19.3) | 1528 (56.1) | 5 (5.2) | 7 (19.4) | <0.001 |
| **Any Surgery Trial, N (%)** | 47 (4.3) | 113 (5.2) | 10 (2.3) | 1 (1.8) | 363 (13.3) | 1 (1.0) | 0 (0.0) | <0.001 |
| **Only Multicenter Trials, N (%)** | 993 (90.7) | 2142 (97.7) | 428 (98.6) | 46 (80.7) | 2688 (98.7) | 90 (93.8) | 36 (100.0) | <0.001 |
| **Only Multinational Trials, N (%)** | 612 (55.9) | 1272 (58.0) | 415 (95.6) | 46 (80.7) | 896 (32.9) | 90 (93.8) | 36 (100.0) | <0.001 |
| **Only Multiregional Trials, N (%)** | 516 (47.1) | 969 (44.2) | 415 (95.6) | 46 (80.7) | 730 (26.8) | 90 (93.8) | 36 (100.0) | <0.001 |
| **Only Industry-Sponsored Trials, N (%)** | 607 (55.4) | 838 (38.2) | 393 (90.6) | 39 (68.4) | 570 (20.9) | 89 (92.7) | 36 (100.0) | <0.001 |
| **Only Systemic Therapy Trials, N (%)** | 714 (65.2) | 1431 (65.3) | 372 (85.7) | 34 (59.6) | 801 (29.4) | 87 (90.6) | 29 (80.6) | <0.001 |

**FIGURE 1.**
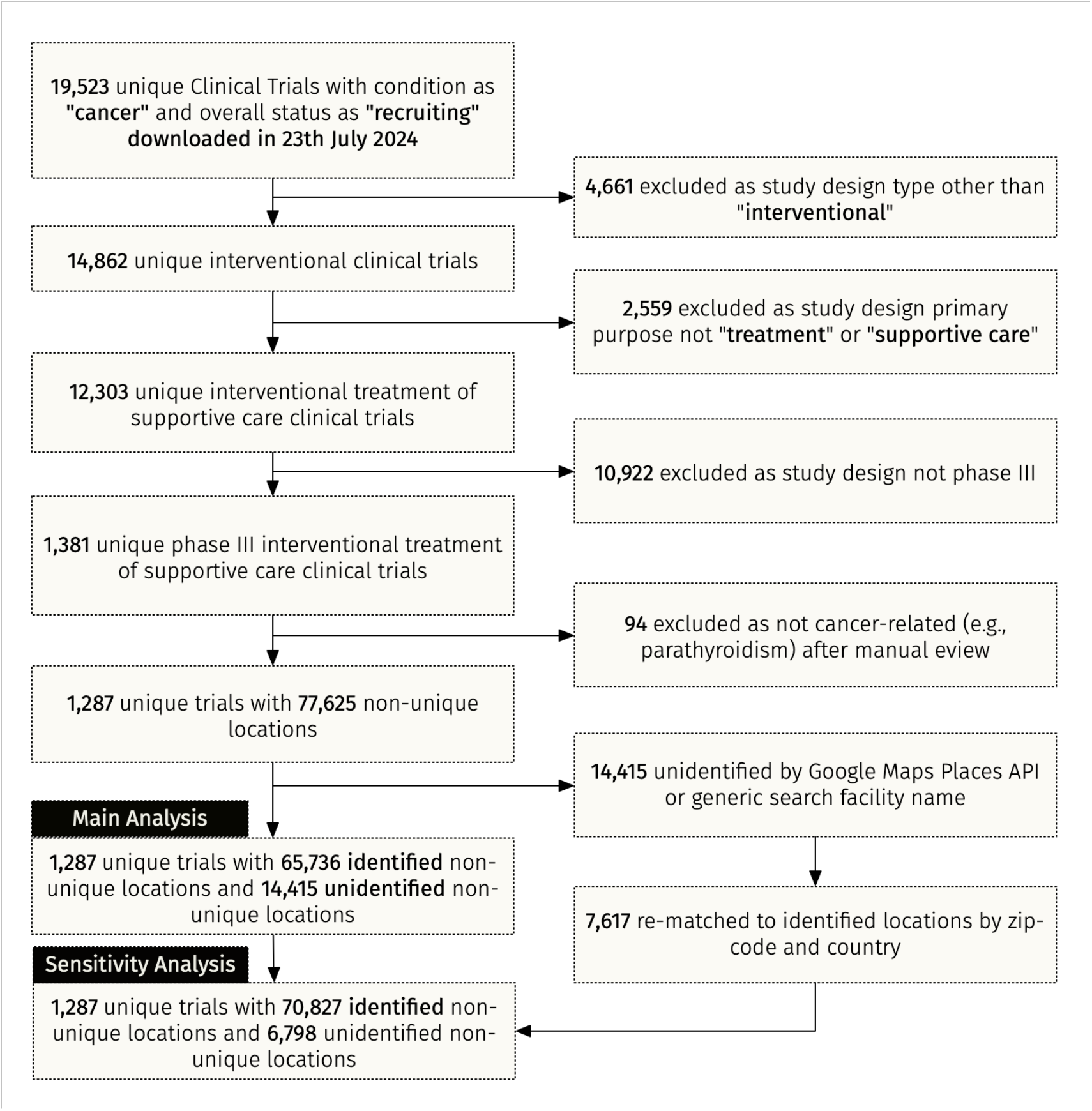
Fluxogram Demonstrating the Database Search Strategy, Eligibility Criteria, and Final Matched Included Studies and Facilities for the Main and Sensitivity Analysis

### Research Facilities Distribution and Characteristics

Of 77,625 non-unique research sites, 65,736 (84.7%) were mapped to 6,634 unique research facilities across 84 countries (**Suppl. Figure 1**). Most research facilities were in the United States (2,626, 39.6%) followed by China (464, 7.0%), Germany (383, 5.8%), France (273, 4.1%) and Japan (263, 4.0%) (**Table 1)**. The median (IQR) number of listed trials per facility was 4 (1-12), with the majority of facilities having 1-5 clinical trials (58.4%), and a minority 50 or more (1.9%). Most research facilities were found in HIC or UPMC (97.7%). Only 3.2% (211) had any single-center trials; and nearly 40% of research facilities had only industry-sponsored trials. LAC (6.5%), SAR (1.4%), MENA (0.9%) and SSA (0.5%) had the lowest share of facilities. **Figure 2** shows the facilities distribution worldwide.

**FIGURE 2.**
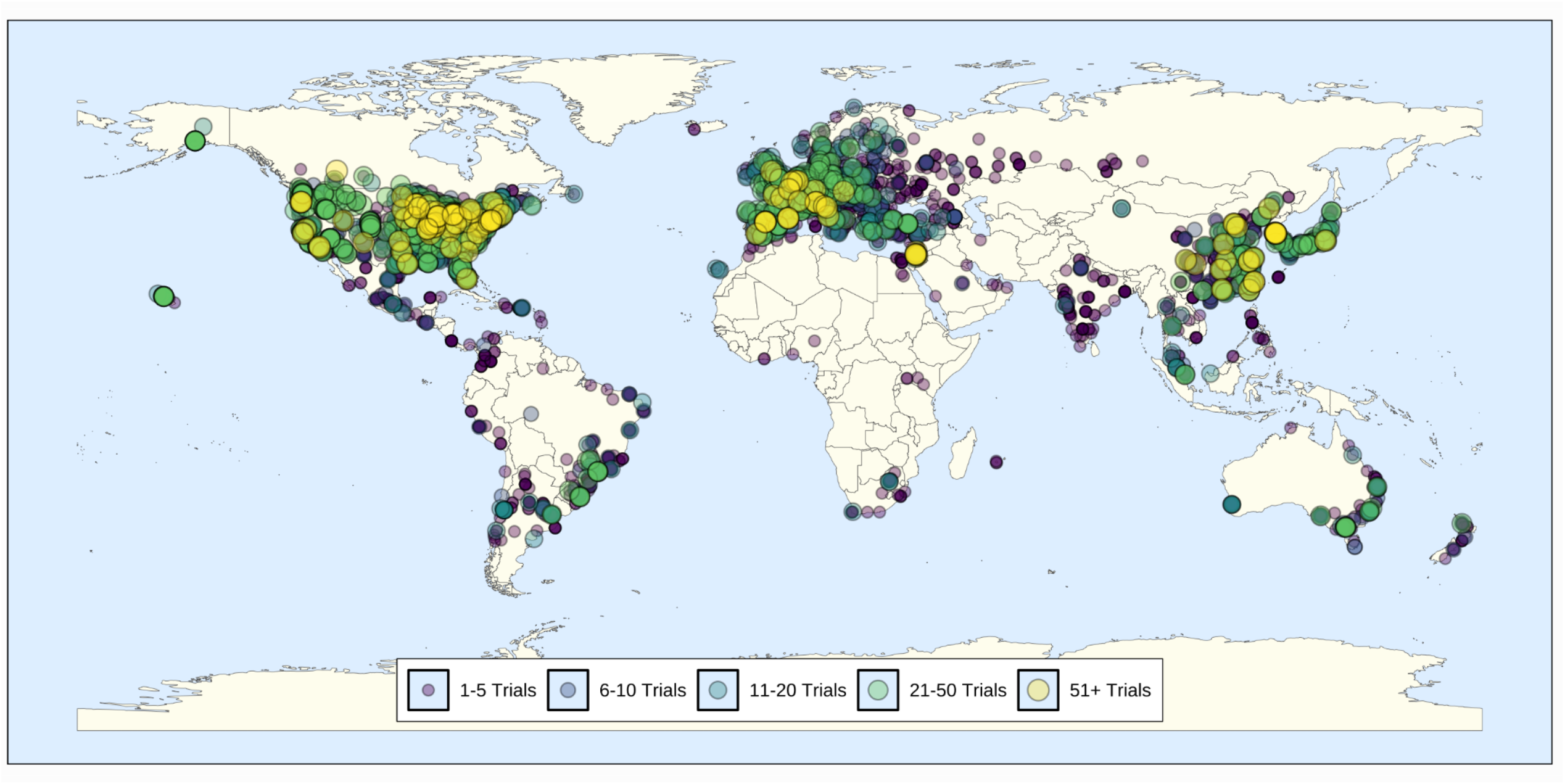
Global Distribution of Research Facilities and Their Respective Size According to the Number of Phase III Cancer Clinical Trials.

Table 2 shows research facilities characteristics across WDI regions. While NAM had larger research facilities with nearly half with 10 or more listed trials, SAR had smaller facilities with most with less than 5 trials (94.8%). Considering research facilities density (per million), NAM had the highest density followed by ECA and EAP. Of the 20 countries with the highest research facilities density, 16 (80%) were from ECA (**Suppl. Figure 2**). LAC, SAR and SSA had the largest proportion of facilities running only multinational (>93%), multiregional (>93%), and industry-sponsored trials (>90%), with all the trials in SSA being industry-sponsored. The proportion of facilities running only systemic treatments trials varied between regions, from only 30% in NAM to 65% in ECA and EAP, and 86% and 91% in LAC and SSA respectively. Among the top 100 research facilities ranked by number of listed trials, the median (IQR) number of listed trials was 62 (57-73.25), and the majority were in the United States (N=53), followed by China (N=9), South Korea (N=7), Spain (N=7), France (N=5), Italy (N=5), and Israel (N=4). All facilities were from HIC or China (**Suppl. Table 5**).

### Association Between Research Facilities and phase III Cancer Clinical Trials

Figure 3 shows the absolute number of cancer trials and research facilities by country on both original and log-transformed scales. China and the United States had the largest number of trials and research facilities respectively, outperforming all other countries. As expected, linear regression on the original scale showed poor fit (R^2^ = 0.40) and non-normal distributed residuals (Shapiro-wilk, p-value<0.001), whereas log-log transformation revealed an approximately linear relationship with improved fit (R^2^ = 0.86, p-value < 0.001). The estimated elasticity coefficient was 0.95 (95% CI: 0.87 - 1.04), indicating that each 1% increase in research facilities was associated with a 0.95% increase in available phase III cancer trials. Model diagnostics showed no evidence of heteroskedasticity (Breusch-Pagan, p-value=0.27) and residuals were approximately normally distributed (Shapiro-wilk, p-value=0.43). Linear regression diagnostics showed the United States as an outlier and its removal yielded overall similar estimates (R^2^ = 0.87, coefficient 1.00 95% CI 0.92 - 1.09).

### Correlation Between Research Facilities and WDI Indicators

We found strong significant correlations between the number of research facilities and GDP value (r = 0.92 and ρ = 0.79, p<0.001), and moderate strength correlations between health expenditure (r = 0.50 and ρ = 0.52, p<0.001), research expenditure (r = 0.52 and ρ = 0.51, p<0.001), and total population (r = 0.58 and ρ = 0.55, p<0.001), with the remaining having weak correlation (**Suppl. Figure 3)**.

**FIGURE 3.**
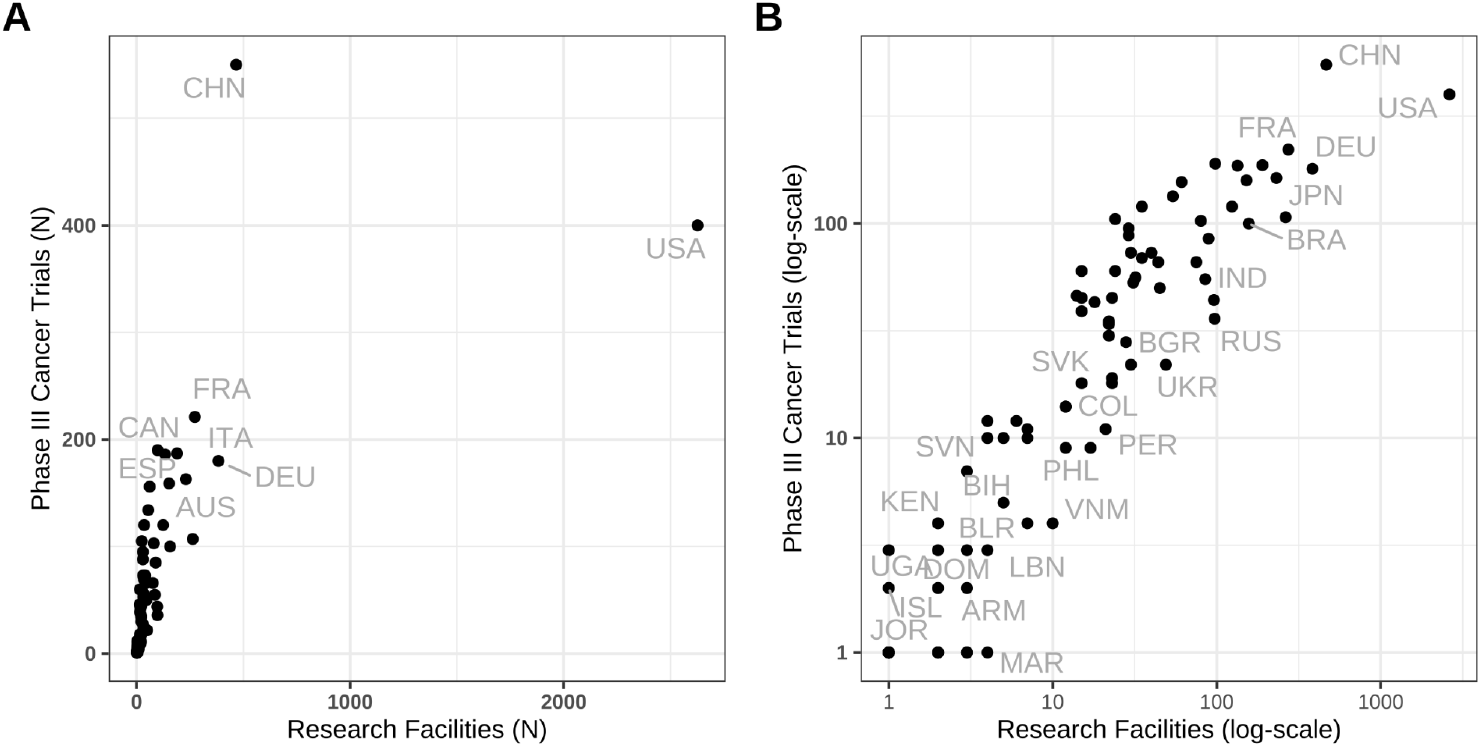
Scatter plot of number of phase III clinical trials (y-axis) and number of research facilities (x-axis) in the original (A) and log-scale (B) by country.

### Sensitivity Analysis

Re-identification of locations using the unique zip-code and country increased the proportion of identified locations to 70,827 (91,24%) (**Suppl. Figure 1**). We observed an increase in the size of research facilities across all regions, more pronounced in LAC, SAR and ECA (**Suppl. Figure 4**). These findings were reflected in a higher proportion of countries from EAP (N=25), ECA (N=24) and MENA (N=5) among the largest 100 research facilities in size with a corresponding decrease in NAM (N=46) (**Suppl. Table 6**). However, interpretation of differences across regions (**Suppl. Table 7)** remained similar. The elasticity coefficient between phase 3 cancer trials and research facilities slightly increased (elasticity coefficient 1.00, 95%CI 0.91 - 1.09, R^2^ = 0.86, p-value < 0.001) (**Suppl. Figure 5)**.

### Unidentified Locations Correlates

We observed a higher proportion of unidentified locations for LAC, India, Egypt and Eastern Europe, decreased when we re-identified locations by zip-code in the sensitivity analysis (**Suppl. Figure 7)**. For the main analysis, multilevel logistic regression revealed a significant regional disparity in location identification (NAM as the reference) between WDI regions (**Suppl. Table 8)**: LAC has the highest odds of unidentification (OR 4.46 95%CI 3.82–5.21), followed by ECA (OR 2.08 95%CI 1.84–2.36) and SAR (OR 2.37 95%CI 1.60–3.53). Industry-sponsored trials were nearly 15 times more likely to have unidentified locations compared to government-sponsored trials (OR 15.22 95%CI 5.35–43.35). Estimates remained similar in the multivariable model. After facilities re-identification in the sensitivity analysis, the odds of unidentified location decreased for LAC (OR 1.52 95%CI 1.34–1.72), and were no longer significant for ECA (OR 0.91 95%CI 0.83–0.99). However, industry-sponsored trials remained highly associated with unidentification of facilities (OR 10.39 95%CI 4.74–22.76).

## DISCUSSION

We have mapped the global distribution and characteristics of research facilities conducting phase III cancer clinical trials. Our findings show that regions predominantly comprising high-income countries have a greater number and higher density of research facilities, which are also larger in scale and less reliant on industry, compared to regions with predominantly lower-income countries. Interestingly, we have shown that the number of research facilities within a country is highly associated with the number of phase III clinical trials available, in an approximately 1:1 log-log relationship, meaning a 1% increase in research facilities correlates with a nearly 1% increase in phase III clinical trials availability. Although the direction of the correlation is expected, this unitary (1:1) constant returns to scale mirrors the proportional elasticity of investment often seen in classical economic models^31^, suggesting that trial capacity is directly limited by, and responsive to, physical institutional growth. This finding can help policymakers plan research infrastructure investment.

Our analysis shows that research infrastructure is well-developed among HIC, particularly in North America and European countries. The US holds the highest absolute and second highest density estimates of research facilities globally, including nearly half of the world’s largest facilities with the highest trial volume, while European and Central Asian nations comprise 80% of the top 20 countries with the highest research facilities density. This extensive infrastructure likely contributes to the elevated density of cancer trials and the observed independence from industry influence reported in numerous studies^23,25,27,32^. Among UMIC and LIC countries, China represents a notable exception. Despite a research infrastructure less than five times that of the US, China has emerged as the leader in the number of available phase III cancer trials, characterized by a high proportion of single-center studies. This observation likely suggests a strategic shift in Chinese investment toward larger, multi-institutional organizations possessing the capacity to conduct concurrently multiple phase III clinical trials. Consequently, China’s pharmaceutical landscape is witnessing an increasing number of drug approvals, both domestically^33^ and internationally^34^.

In developing regions, cancer research remains largely constrained by industry-sponsored, multiregional systemic trials conducted within a research infrastructure characterized by limited alternative options. For example, LAC and SSA have 0% and 1.4% of their research facilities conducting single-center phase III clinical trials with a proportion greater than 90% of them conducting only industry-sponsored trials. This may be partially attributed to the high prevalence of small research facilities (≤ 5 listed trials) exceeding 70%; however, the more significant contributors appear to be chronic deficiencies in governmental research funding, coupled with limited human resources and persistent operational bureaucracies^28,35^. Recent efforts by regional oncology cooperative groups to design independent trials are ongoing, but their impact remains to be determined^35^. However, from an optimistic standpoint, existing research infrastructure within these regions offers a foundation for future progress, suggesting that strategic goals should prioritize capacity building and increased funding for independent research planning.

ClinicalTrials.gov is the most comprehensive database of registered clinical trials in absolute numbers and in available data granularity concerning participating research facilities, which makes it useful not only for research purposes^24,25,36^ but also for updated information of available clinical trials from the provider perspective^5^. Our analysis revealed that approximately 15% of location records, or 10% following zip code re-matching in sensitivity analysis, remained unidentifiable, with a disproportionate amount attributable to industry-sponsored imputed data and locations from developing regions. This can be partially explained by Google Maps coverage limitations^29^, but even in regions where Google Maps have extensive coverage as LAC, the proportion of unidentifiable locations remained relatively high compared to other countries. Developing regions had a higher proportion of research facilities with address-mapping inconsistencies, potentially reflecting lower prioritization or reduced data curation efforts for these sites within ClinicalTrials.gov. While complete and identifiable location information is not a regulatory requirement for FDA approval, its encouragement is warranted given the database’s widespread utility as a source of study availability.

This study has many strengths. This is the first study to provide a comprehensive mapping of research infra-structure conducting phase III cancer trials worldwide, with results that can directly impact national policies in terms of research expenditure and capacity investments. We have adopted a data-driven approach and leveraged large language models (*Gemini*) to help identify research facilities using a hybrid human-AI approach. In fact, the use of *Gemini* to cluster facilities within the same location overcame potential limitations of geographic knowledge introduced by the authors and provided a more unbiased estimate. This geospatial analysis demonstrated Google Maps’ validity to accurately identify and delineate study sites on a global scale, suggesting broader applications in real-world applications including epidemiological surveillance and resource allocation assessments.

This analysis is subject to limitations. First, the data utilized were restricted to those available on ClinicalTrials.Gov, potentially underestimating the total number of trials conducted globally^37,38^. The WHO database of clinical trials, which aggregates from diverse sources, provides a more comprehensive coverage of clinical trials available, but does not report sufficient granular data for facilities identification^39^. Second, identification of research facilities relied on Google Maps data, which may have been incomplete due to insufficient ClinicalTrials.Gov information, lack of facility registration, or limited geographic coverage within certain countries^29^. To address this, a sensitivity analysis was performed, re-identifying locations using zip-code data, which increased facility identification success to 90%. However, the proportion of unidentifiable locations remained high in some countries and results should be interpreted in light of these findings. Analysis of Gemini-Pro 3.0 same facilities evaluation demonstrated high accuracy within a subset of Brazilian data (>= 97%); however, performance may vary across countries, with potentially superior accuracy in regions where Google search engine usage is prevalent and reduced accuracy where Google’s coverage is limited. Importantly, the use of Gemini-Pro 3.0 was applied after manual review of the Google Maps results by the authors, and therefore errors may imply under- or overestimation of facilities’ size (because of clustering) but would not affect the facilities’ identification per se. Lastly, this study focused exclusively on phase III trials, and, while these trials represent the largest coverage of research facilities^24,40^ and are critical prior to drug approval, these findings may not be generalizable to other study designs (e.g. phase I or phase II).

In conclusion, our analysis reveals a robust, approximately 1:1 log-log correlation between the number of research facilities and the volume of phase III cancer trials conducted, which has major implications for policymakers planning research infra-structure and national investments in cancer science. The US currently possesses the most extensive research infrastructure globally, while China exhibits a leading number of phase III trials, a consequence likely driven by prior investments in large-size facilities and accelerated drug development programs. Across UMIC and LMIC, research capacity remains substantially constrained, largely focused on systemic therapies and frequently reliant on industry-sponsored, multiregional phase III trials. Therefore, immediate action is needed through targeted capacity-building initiatives and increased investment, with future policy strategies prioritizing the training of local research personnel and the strategic allocation of funding to support independent, locally-driven research initiatives.

## Data Availability

All data used in this study are publicly available from ClinicalTrials.gov. The datasets generated and/or analysed during the current study will be made available by the corresponding author after peer-reviewed publication.

## Contributors

FLN and JMM conceived the study. FLN, RTC, AFV, and FL participated in data collection and validation. FLN led the statistical analysis. FLN, JWR, RTC, AFV, FL and FYM curated and validated the data. FLN wrote the first draft of the manuscript. FYM, JWR, and JMM provided supervision. All authors contributed to and approved the final manuscript. FLN and JMM had full access to and verified the data. JMM acts as the guarantor. The corresponding author attests that all listed authors meet authorship criteria and that no others meeting the criteria have been omitted.

## Data sharing

All data used in this study are publicly available from ClinicalTrials.Gov. The datasets generated and/or analysed during the current study will be made available by the corresponding author after peer-reviewed publication.

## Declaration of Interests

JMM reports consulting or advisory roles for Janssen Oncology and Astellas Pharma; speakers’ bureau participation for Janssen Oncology, Amgen, Zodiac Pharma, Bayer, Astellas Pharma, Ipsen, Pfizer, AstraZeneca, Merck, and Novartis; travel, accommodations, or expenses from Zodiac Pharma and Janssen Oncology; expert testimony for Bayer, Astellas Pharma, and Adium Pharma; honoraria from Janssen, Zodiac Pharma, Astellas Pharma, Bayer, Ipsen, Pfizer, AstraZeneca, Amgen, Merck, and Novartis; and research funding from Bayer. These institutions had no role in the study design, data collection, data analysis, data interpretation, or writing of the report. The remaining authors have no conflicts of interest to disclose.

## Acknowledgements

No external funding was provided for this study. Artificial intelligence based methods were used as part of the study methodology, as described in the Methods section. During the preparation of this manuscript, AI-assisted tools (Gemini-Flash 2.5, Gemini-Pro 3.0) were employed for proofreading to correct grammar, spelling, syntax, and clarity. No generative AI or AI-assisted technologies were used to generate the scientific content. The text has been reviewed, edited, and approved by all authors, who take full responsibility for its content.

## SUPPLEMENTARY APPENDIX

### Methodology for Research Facilities Retrieval and Identification

Briefly, we aim to describe and characterize research facilities conducting phase III clinical trials in cancer. We used the ClinicalTrials.gov database because of its comprehensiveness in listing the locations participating in each study. However, the location information is in unstructured format and, therefore, Google Maps (through its application programming interface [API]) was utilized as a standardized identifier of each research site. To account for instances where a single institution might have multiple Google Maps listings, unique Google Maps IDs within 1,000 meters of each other were evaluated as representing the “same facility.” Google Gemini Pro served as an additional reviewer in this step, particularly addressing author geographical knowledge limitations and language barriers; its performance was optimized using prompt engineering against a “gold standard” random sample of locations from Brazil. These identified pairs were then used to group (cluster) research facilities deemed to be identical. The detailed methodology is described below, and associated code is available on GitHub (https://github.com/felippelazar/CancerTrialsFacilitiesGlobalDistribution/).

#### 1 Database Search Strategy

We performed a cross-sectional analysis of all clinical trials with “cancer” as “condition” and “overall status” as “recruiting” from the ClinicalTrials.Gov database using their application programming interface (API) (**https://clinicaltrials.gov/api/v2/studies**) on **23th, July, 2024**. For every trial, we extracted study identification number (NCT), title, eligibility criteria, conditions, design type, design phase, primary purpose, sponsor lead name, sponsor type and all their listed locations, including name, formatted address, city, state, zip code, country and recruitment status. This preliminary database was further **filtered** to include only trials listed as **design “phase III”, design type “interventional”, and primary purpose “treatment” or “supportive care”**. Trial information was manually reviewed, and those with cancer as not the main research focus (e.g., “parathyroidism”) were excluded. **We chose the ClinicalTrials.Gov database given it is the most comprehensive database on clinical trials with available granular data on recruiting locations (e.g. name, city) to potentially identify them**.

#### 2. Creation of Research Facilities Unique Identification

For every listed location in ClinicalTrials.Gov, we created a unique identifier using the combination of its facility name, city, state, zip code and country, after text sanitization. Here is the code snippet used for the creation of the unique identifier:

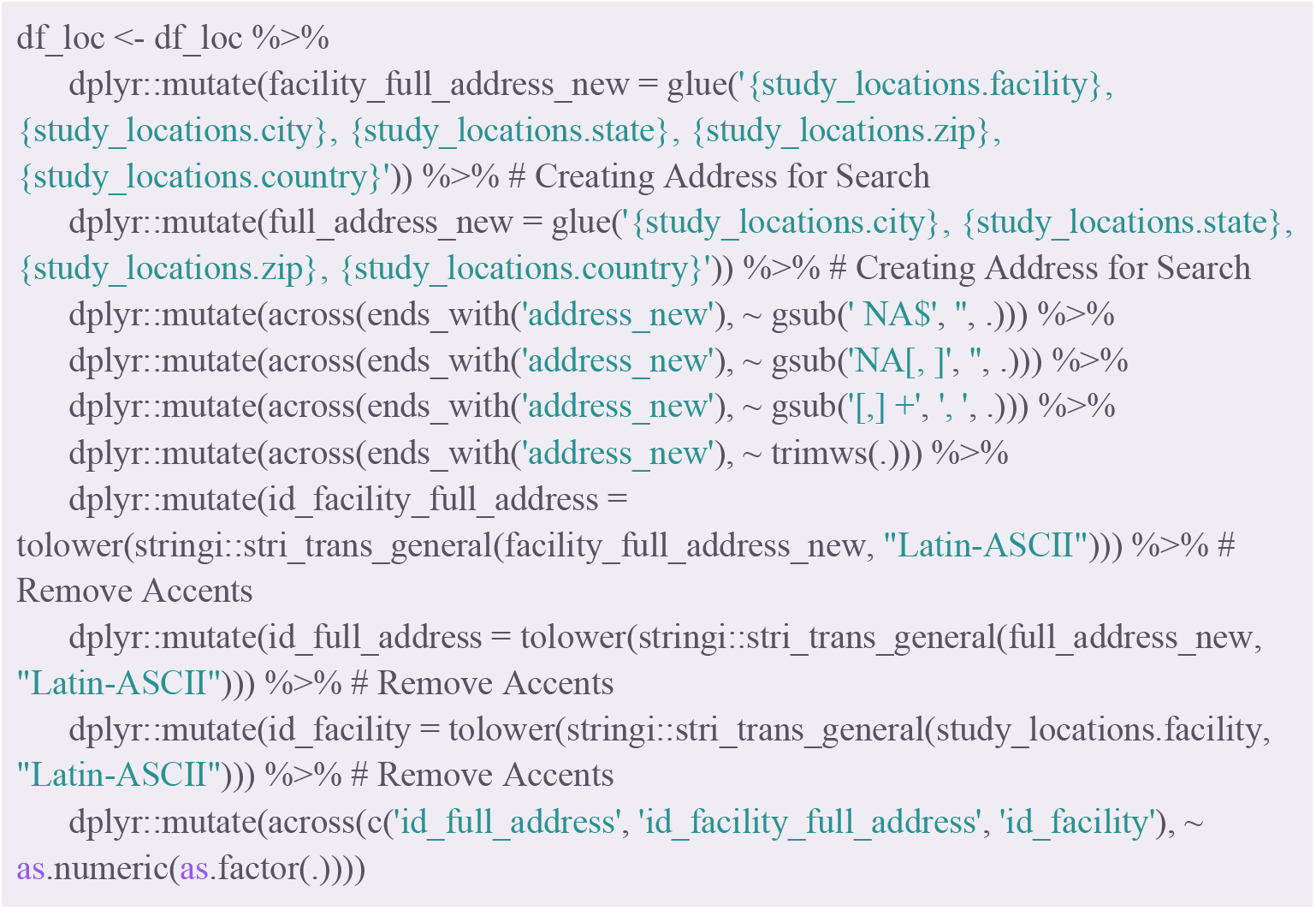

#### 3. Google Maps Query

After duplicate identifiers were removed, unique locations were queried through the Google Maps API (**https://maps.googleapis.com/maps/api/place/findplacefromtext/json)**, using the full address string formatted as ‘facility name, city, state, zip code, country’. For each matched candidate, the API returned candidate places metadata including the Google place identification, the formatted address, country, and geocoordinates (latitude and longitude). **Every pair of unique locations and Google places were manually reviewed by the authors (Suppl. Figure 1)**. Locations where the returned Google places did not correspond to the address location, were determined as unidentified. **Facility locations with generic names were initially classified as already “unidentified” and were not queried through the Google Maps API (below)**.

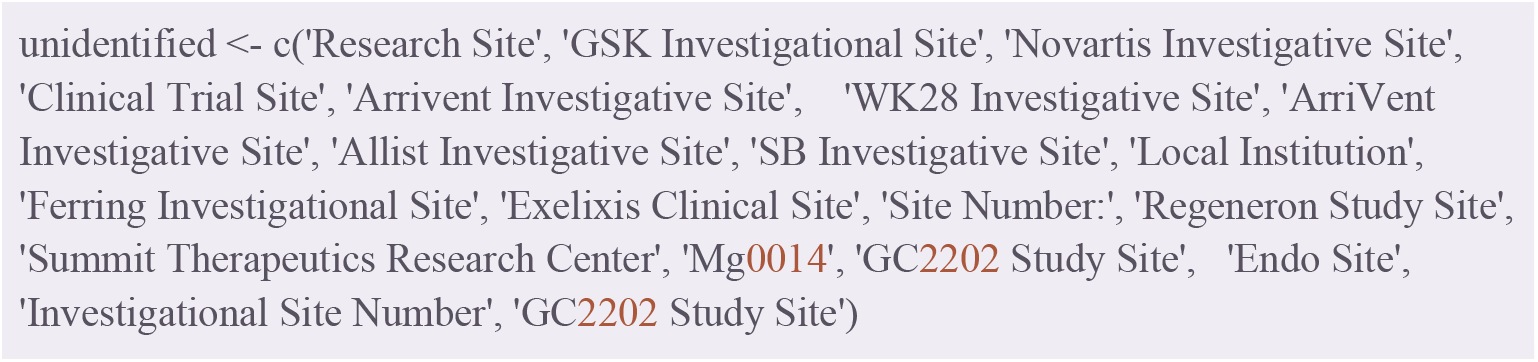

#### 4. Grouping (Clustering) of Same Facility Identifiers

First, **we retrieved all candidate Google addresses IDs within 1**,**000 meters of Haversine distance using their geocoordinates and separated them into pairs** (Example: A and B, B and C, and A and C, if A, B, and C are within 1,000 meters of distance). Second, two authors (AV and RT) manually reviewed each pair of Google addresses and determined if they corresponded to the same research facility. However, because of the highly specific geographic knowledge needed for this step, and language barriers for some institution names (e.g. non-latin alphabet), we used a large language model (LLM) as an additional reviewer. We chose the Gemini Pro because of its performance, thinking capabilities, and its Google search tool calling which allows search of the Google knowledge database to provide answers^1^. During the study period, we used both the Gemini Pro 2.5 and Gemini Pro 3.0, the latter as soon as it became available. We iteratively optimized a prompt against a random sample of 80 location pairs from Brazil, where the authors had sufficient knowledge to provide a gold standard **(Suppl. Table 1)**. We used a **temperature of zero** and a **“high” thinking level**, using the **Google AI Studio (https://aistudio.google.com/)**. The prompt-engineering process improved the accuracy of the output as well as the use of the latest Gemini-Pro model (3.0):

| Model | Prompt Version | Accuracy |
| --- | --- | --- |
| Gemini Pro 2.5 | V1 | 70/80 (N=87.5%) |
| Gemini Pro 2.5 | V2 | 70/80 (N=87.5%) |
| Gemini Pro 2.5 | V3 | 70/80 (N=87.5%) |
| Gemini Pro 2.5 | V4 | 72/80 (N=90.0%) |
| Gemini Pro 3.0 | V4 | 79/80 (N=98.8%) |

We evaluated the output stability by repeatedly querying it with identical prompts (N=10):

| IT | Prompt Version | Wrong IDs | Accuracy | Accuracy Different Facilities | Accuracy Same Facilities |
| --- | --- | --- | --- | --- | --- |
| 1 | V4 | 154 6174 | 78/80 (97.5%) | 45/45 (100%) | 33/35 (94.43%) |
| 2 | V4 | 6653 154 | 78/80 (97.5%) | 45/45 (100%) | 33/35 (94.43%) |
| 3 | V4 | 6175 154 8973 | 77/80 (96.3%) | 44/45 (97.8%) | 33/35 (94.43%) |
| 4 | V4 | 6175 154 | 78/80 (97.5%) | 45/45 (100%) | 33/35 (94.43%) |
| 5 | V4 | 2309 154 | 78/80 (97.5%) | 45/45 (100%) | 33/35 (94.43%) |
| 6 | V4 | 2309 6175 154 | 77/80 (96.3%) | 45/45 (100%) | 32/35 (91.14%) |
| 7 | V4 | 2309 6175<br>6653 154 | 76/80 (95.0%) | 45/45 (100%) | 31/35 (88.6%) |
| 8 | V4 | 154 | 79/80 (98.8%) | 45/45 (100%) | 34/35 (97.1%) |
| 9 | V4 | 6175 6653 153 | 77/80 (96.3%) | 45/45 (100%) | 32/35 (91.14%) |
| 10 | V4 | 2309 6175<br>7255 | 77/80 (96.3%) | 45/45 (100%) | 32/35 (91.14%) |

The final prompt (V4) was applied in all worldwide location pairs. Finally, an additional author (FLN) reviewed and validated the assigned pairs and performed the same research facilities group clustering.

The prompt (V4) used in this study was:

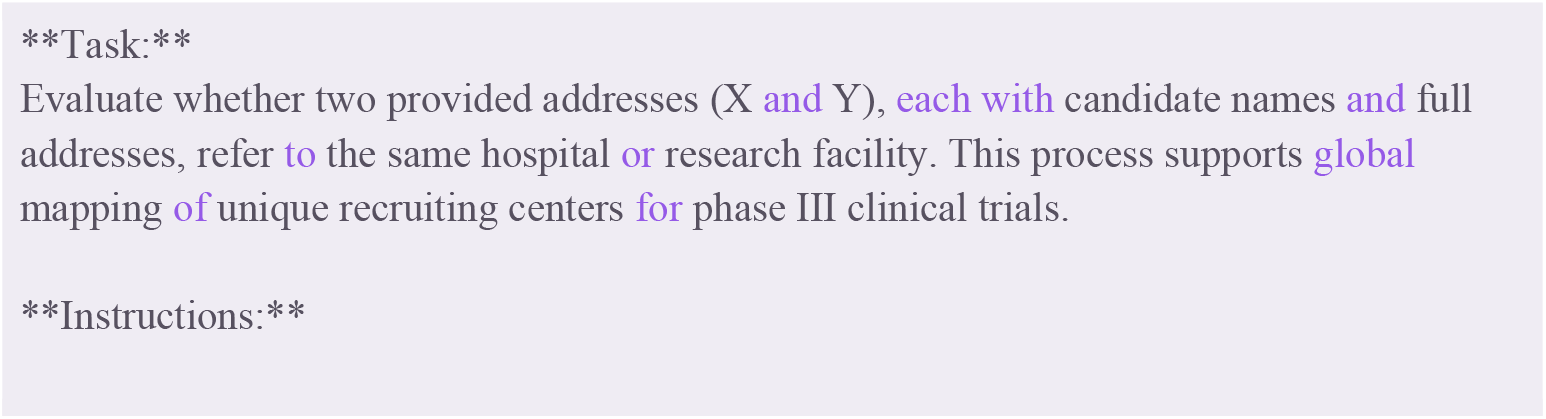
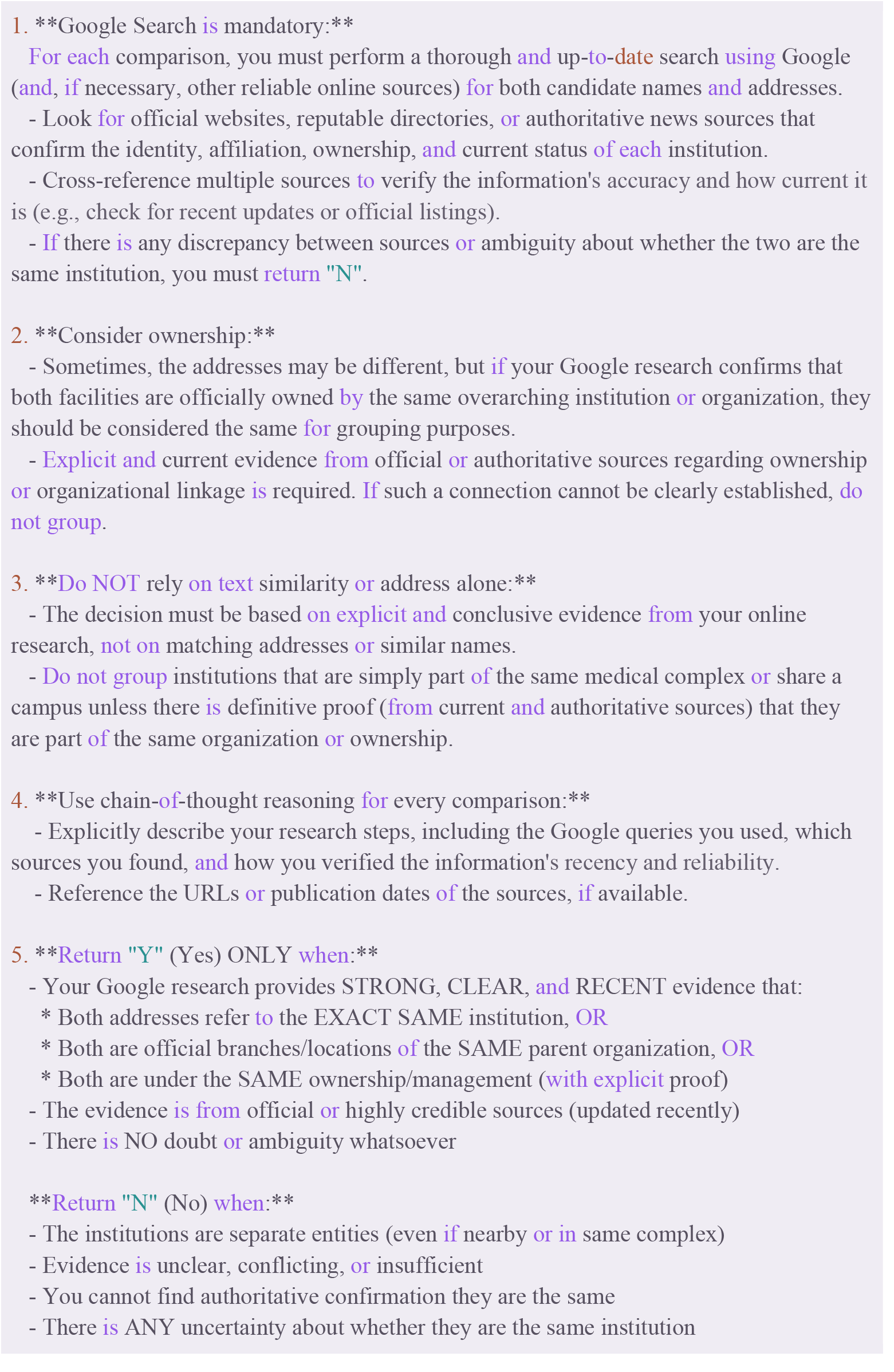
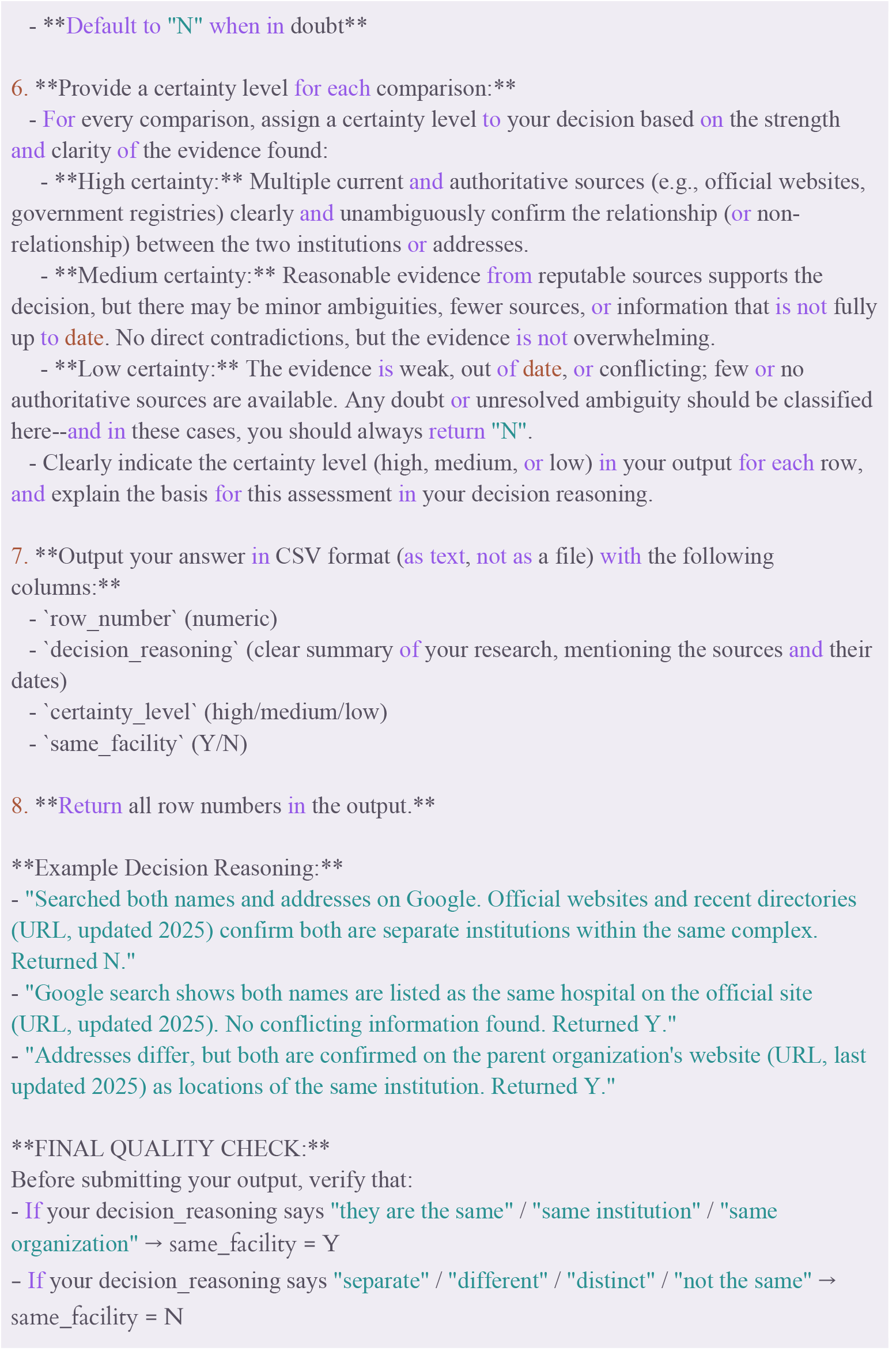
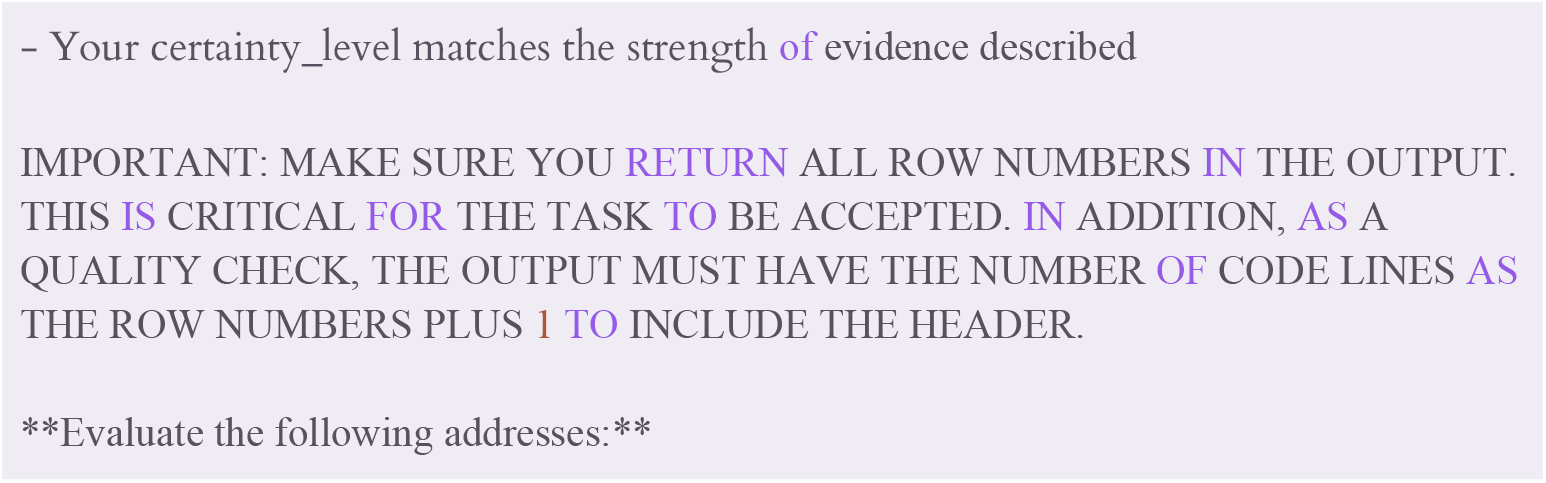

Finally, we performed the same research facilities clustering according to the classified pairs from Gemini Pro 3.0. We chose to use the Gemini Pro 3.0 evaluation rather than the authors’ manual review due to the model’s high accuracy and to minimize potential bias from human evaluation, particularly given the geographic and linguistic limitations inherent in manual review across diverse international institutions.

---

**SUPPL. FIGURE 1.**
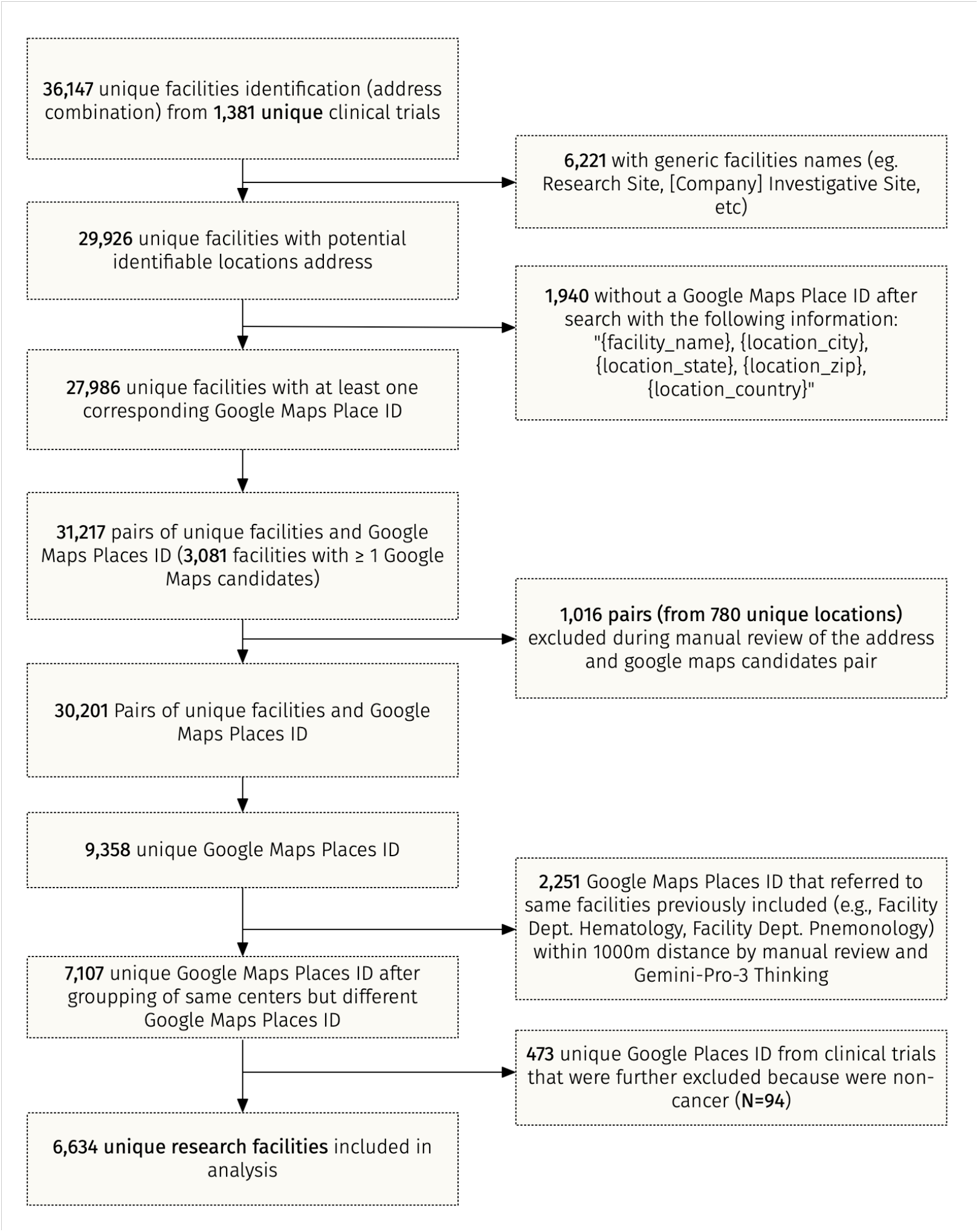
Fluxogram of Research Facilities Identification using Google Maps.

**SUPPL. FIGURE 2.**
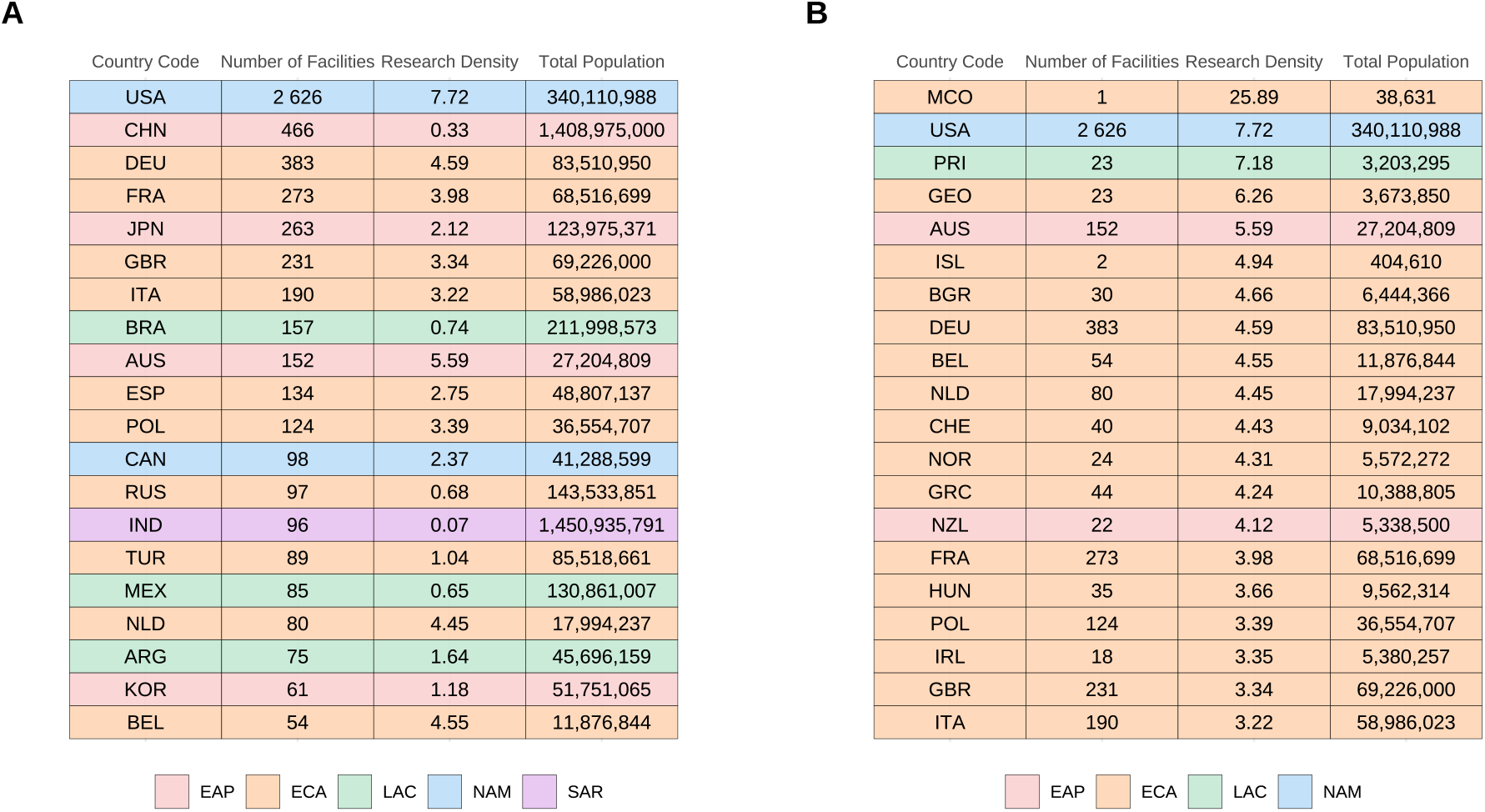
Top 20 Countries by Research Facilities Absolute Number and Density. (A) Top 20 countries ranked by absolute number of research facilities. (B) Top 20 countries ranked by research facility density (number of facilities per million population). Country names are displayed using ISO3 abbreviations. Colors indicate World Bank Development Indicators (WDI) regions: EAP = East Asia & Pacific; ECA = Europe & Central Asia; LAC = Latin America & Caribbean; MENA = Middle East, North Africa, Afghanistan & Pakistan; NAM = North America; SAR = South Asia; SSA = Sub-Saharan Africa.

**SUPPL. FIGURE 3.**
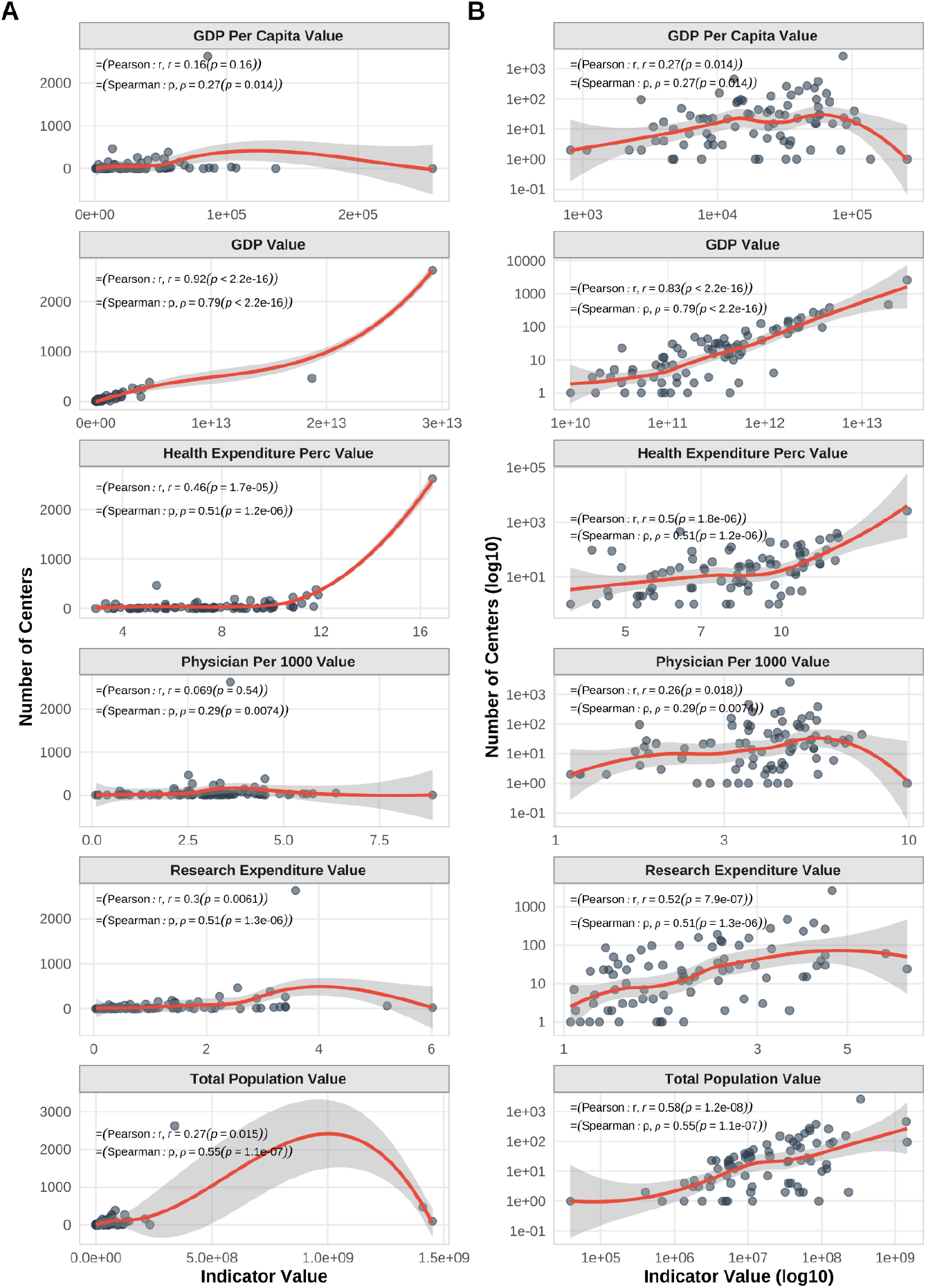
Correlation between Absolute Number of Research Facilities and World Bank Development Indicators. Pearson and Spearman correlation coefficients between absolute number of research facilities and World Bank Development Indicators (WDI) presented in (A) non-log scale and (B) log-scale.

**SUPPL. FIGURE 4.**
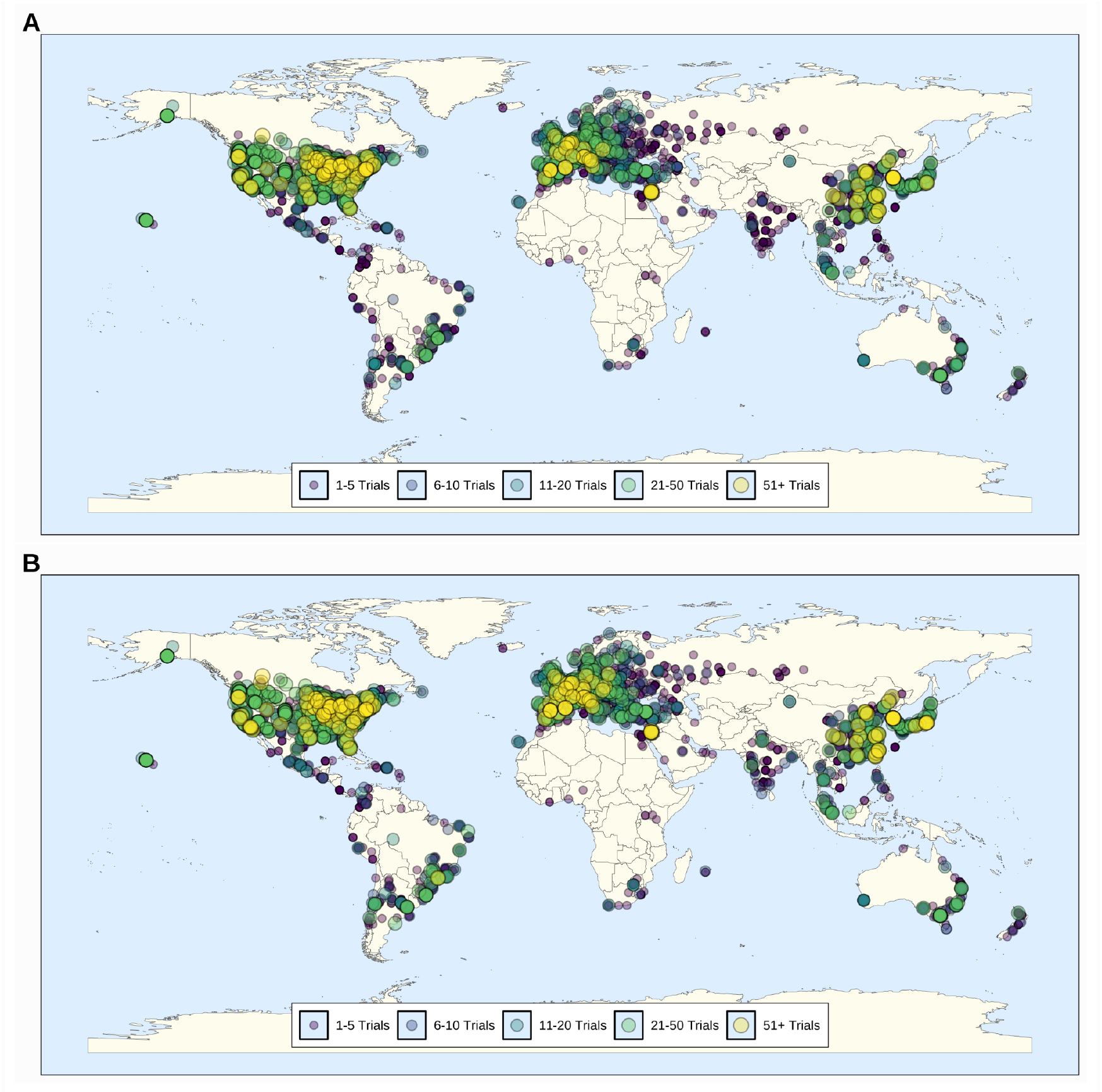
Geographic distribution of research facilities participating in Phase III clinical trials for (A) main analysis and (B) sensitivity analysis. Each circle represents a research facility, with size and color indicating the number of trials conducted (1-5 trials, 6-10 trials, 11-20 trials, 21-50 trials, or 51+ trials).

**SUPPL. FIGURE 5.**
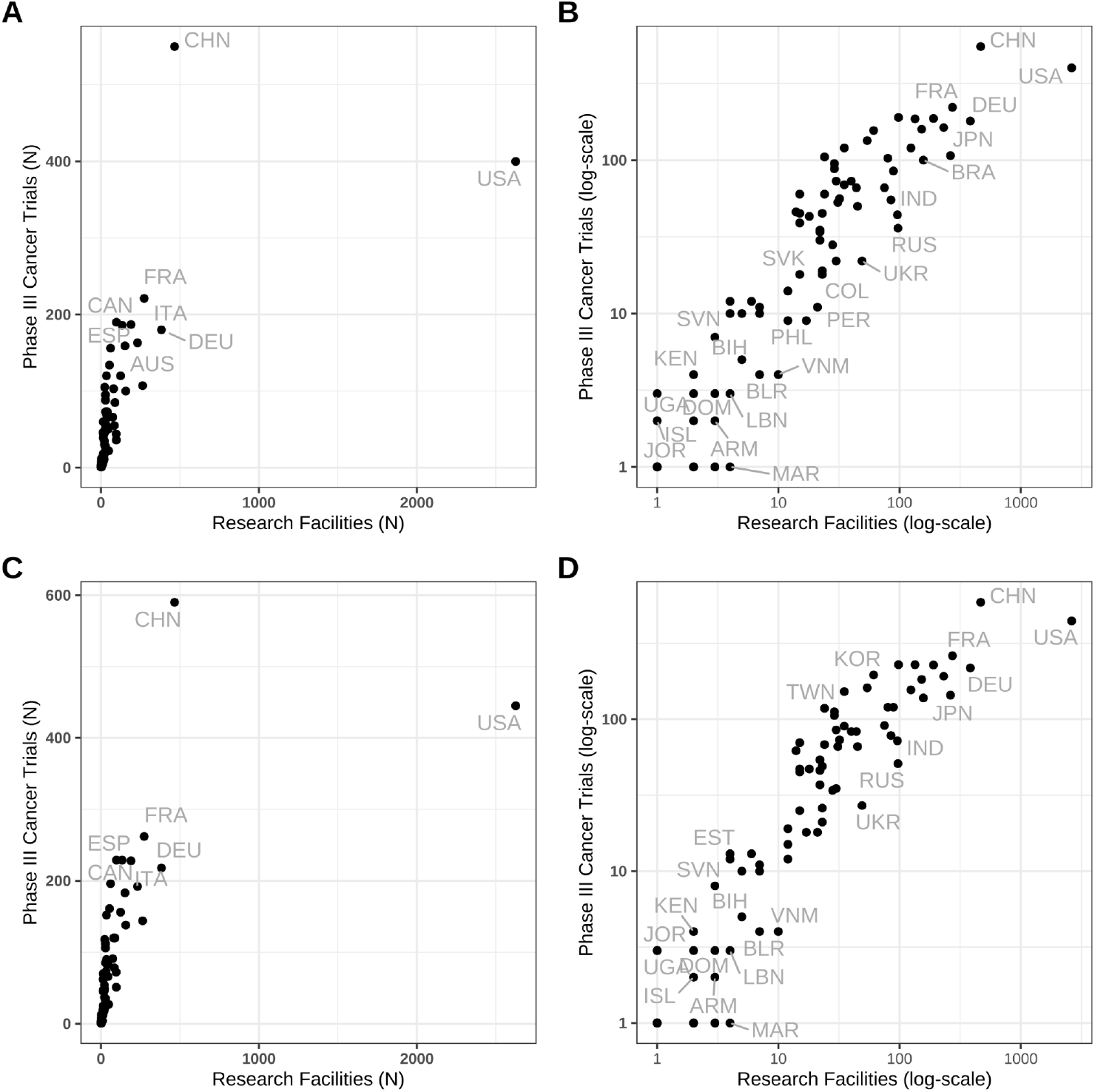
Scatter Plot of Number of Phase III Clinical Trials (y-axis) and Number of Research Facilities (x-axis) in non-log (A and C) and log-scale (B and D) axis by Country for the Main Analysis (A and B) and Sensitivity Analysis (B and C).

**SUPPL. FIGURE 6.**
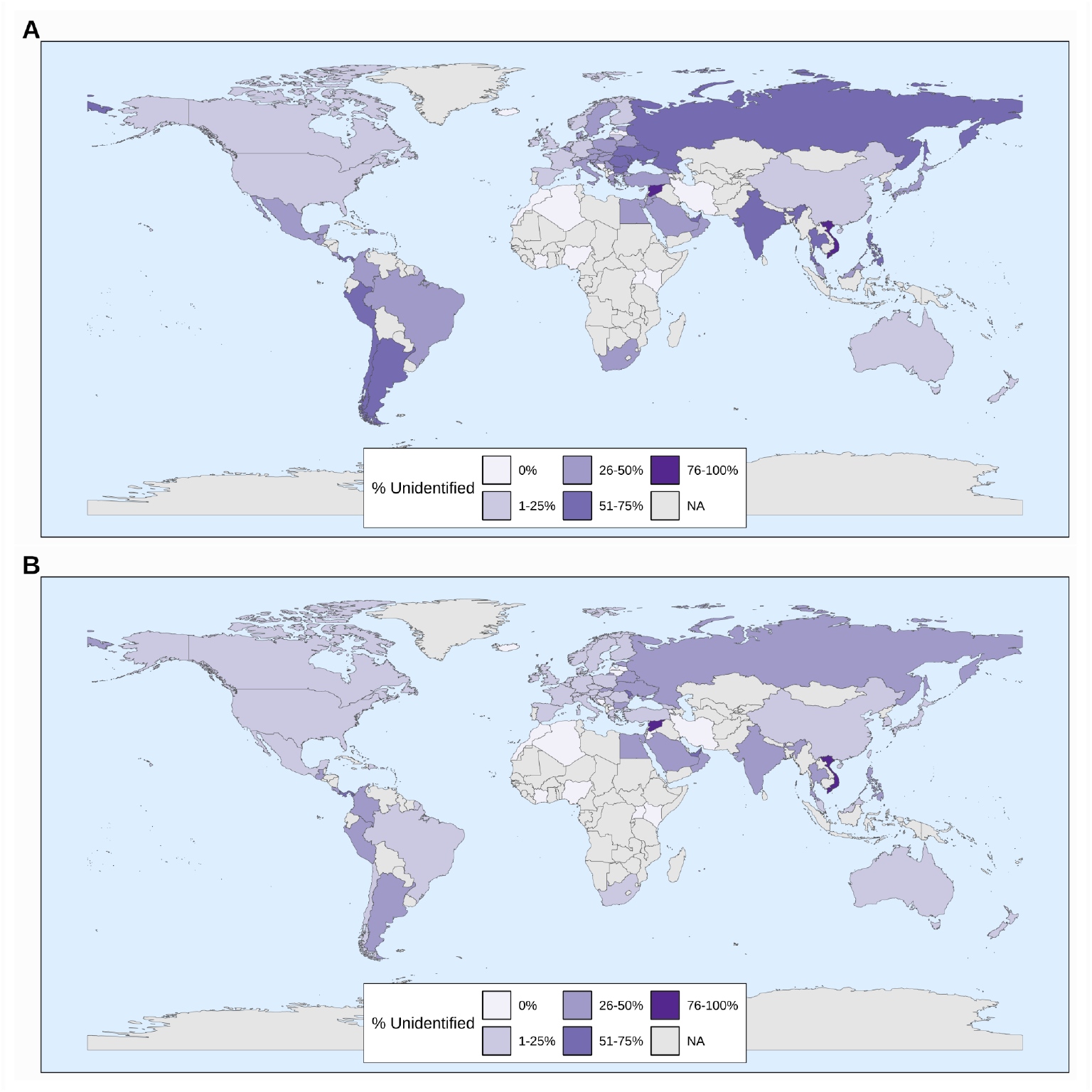
Proportion of Unidentified Locations by Country. Geographic distribution of the proportion of unidentified trial site locations by country for (A) main analysis and (B) sensitivity analysis. Countries are color-coded by percentage of unidentified locations: 0%, 1-25%, 26-50%, 51-75%, 76-100%. NA = not applicable (no trials reported in that country).

**SUPPL. TABLE 1.**
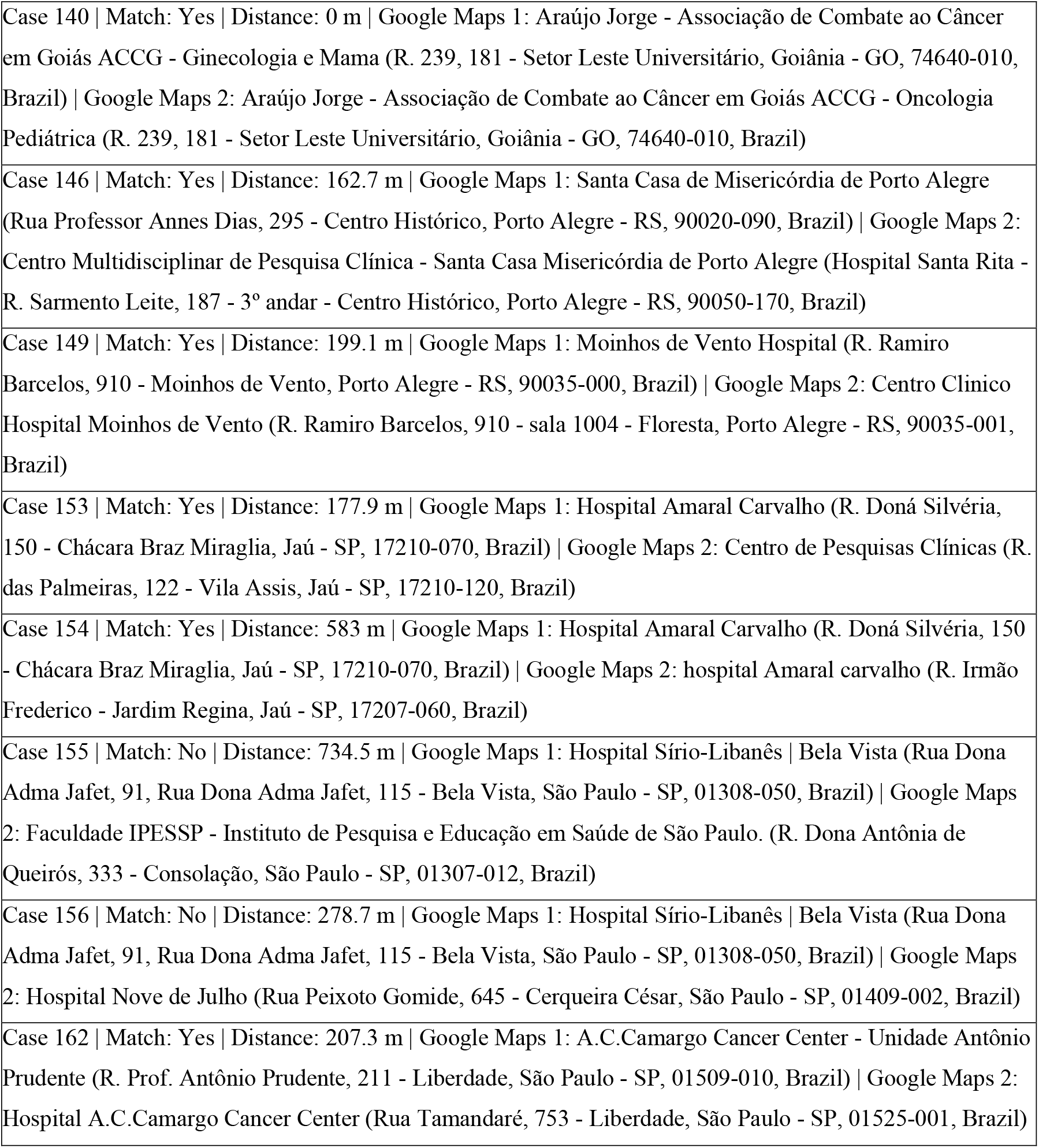
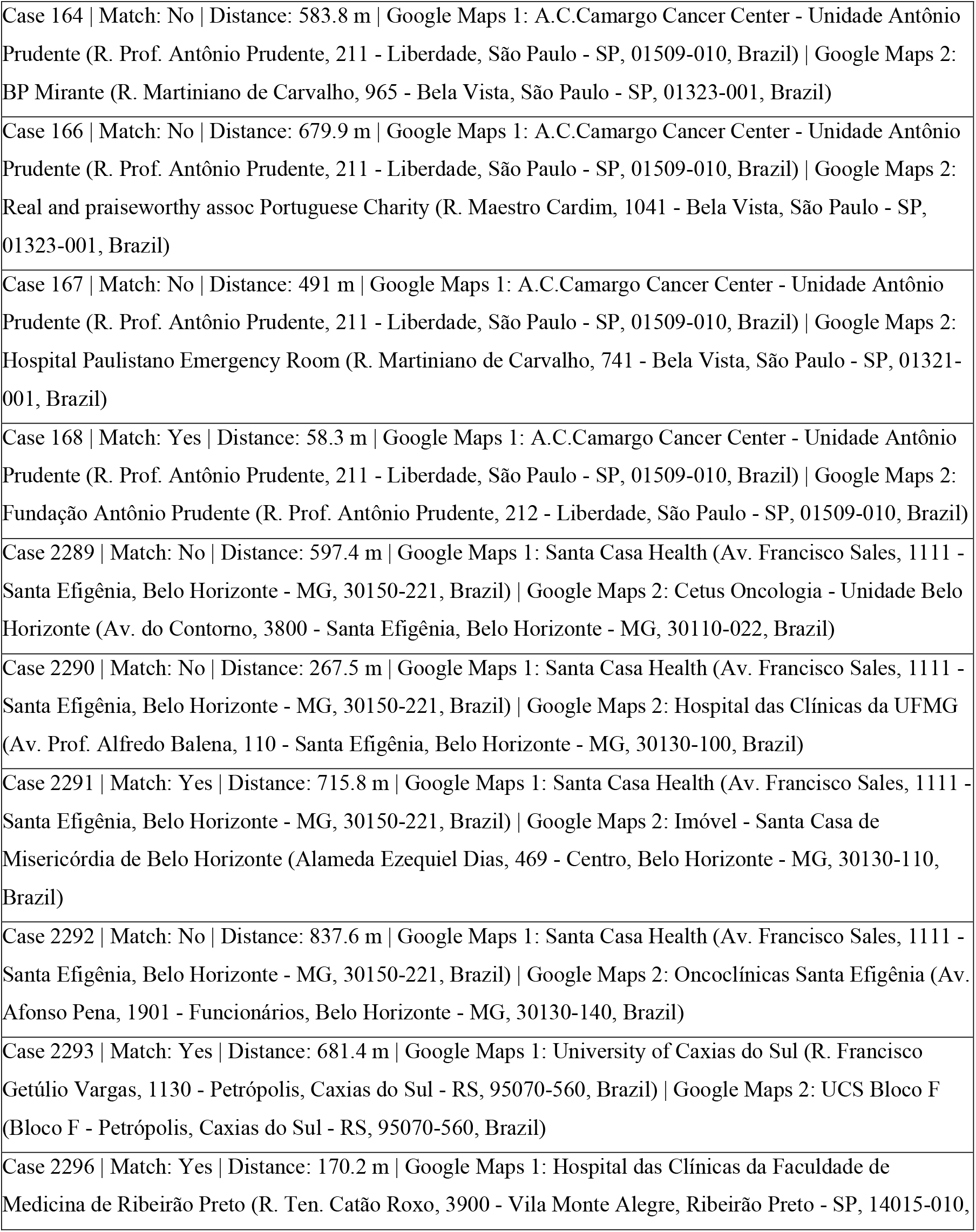
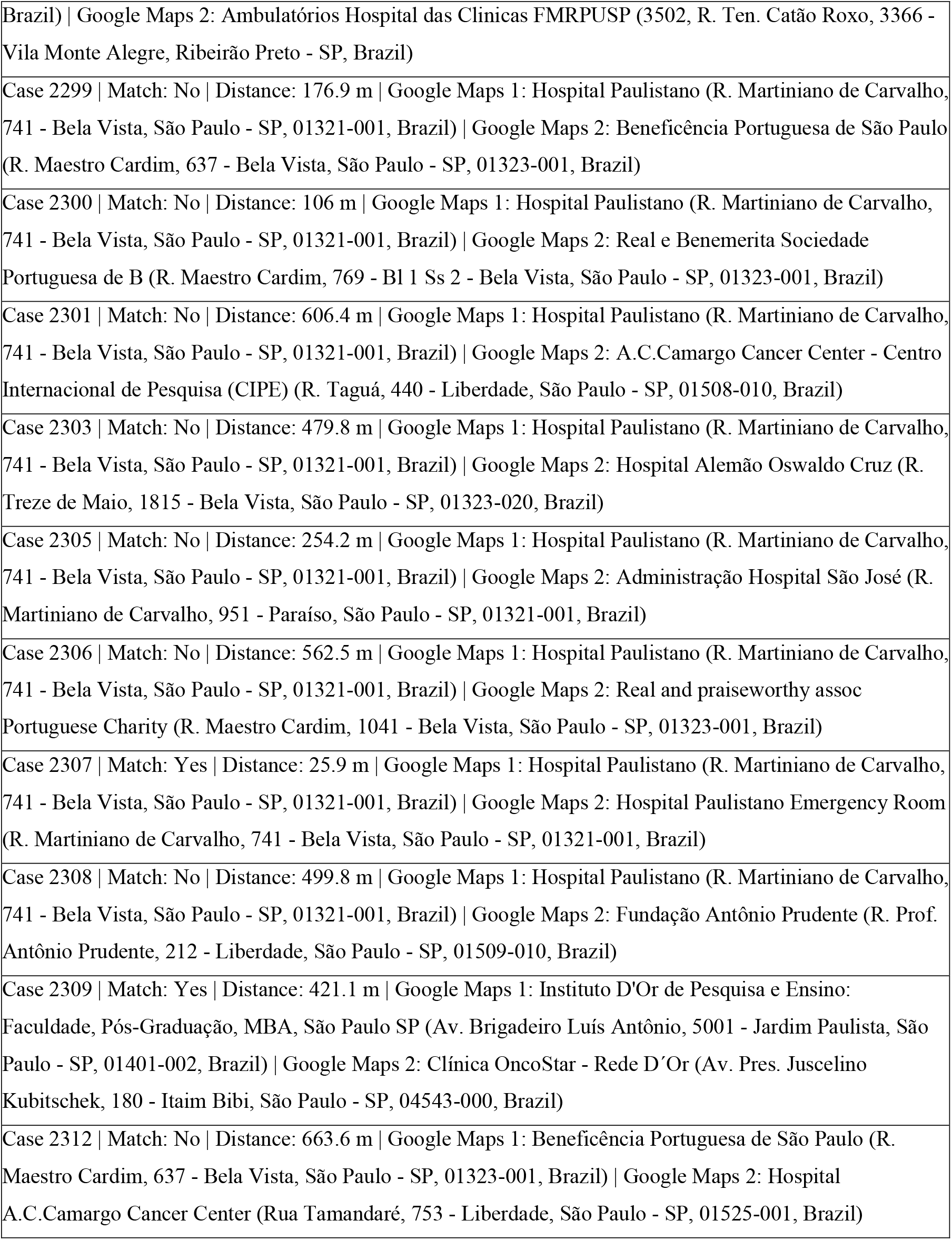
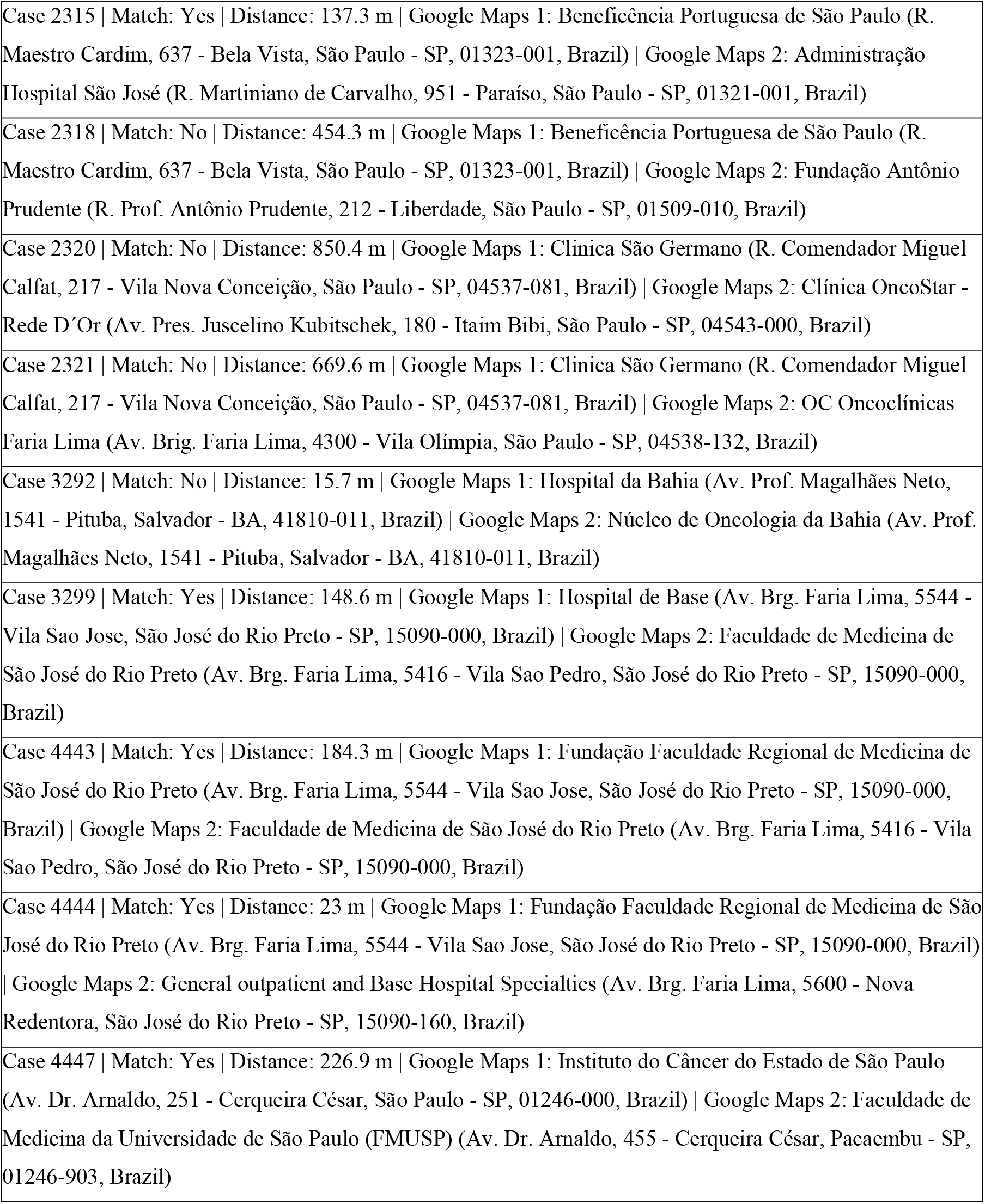
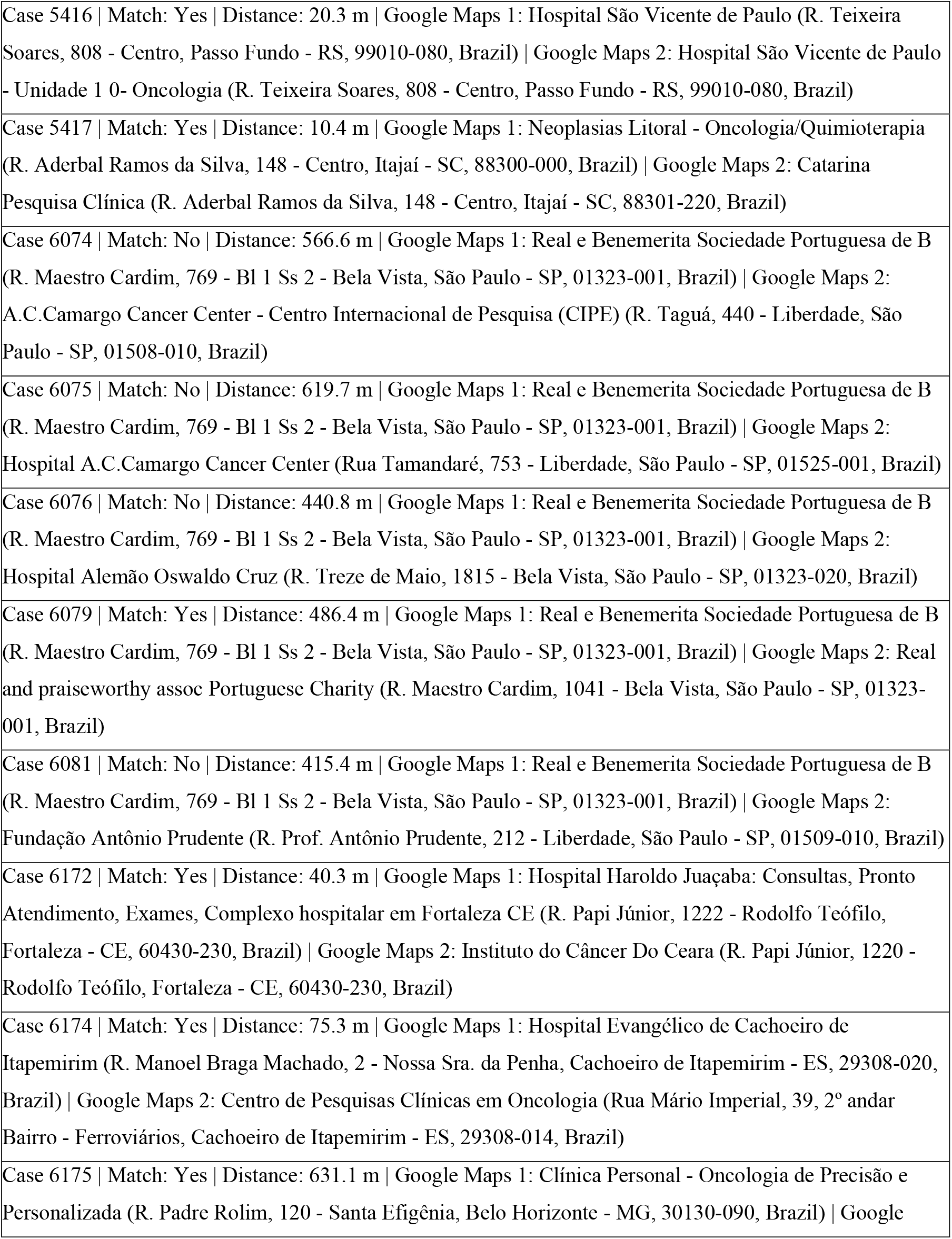
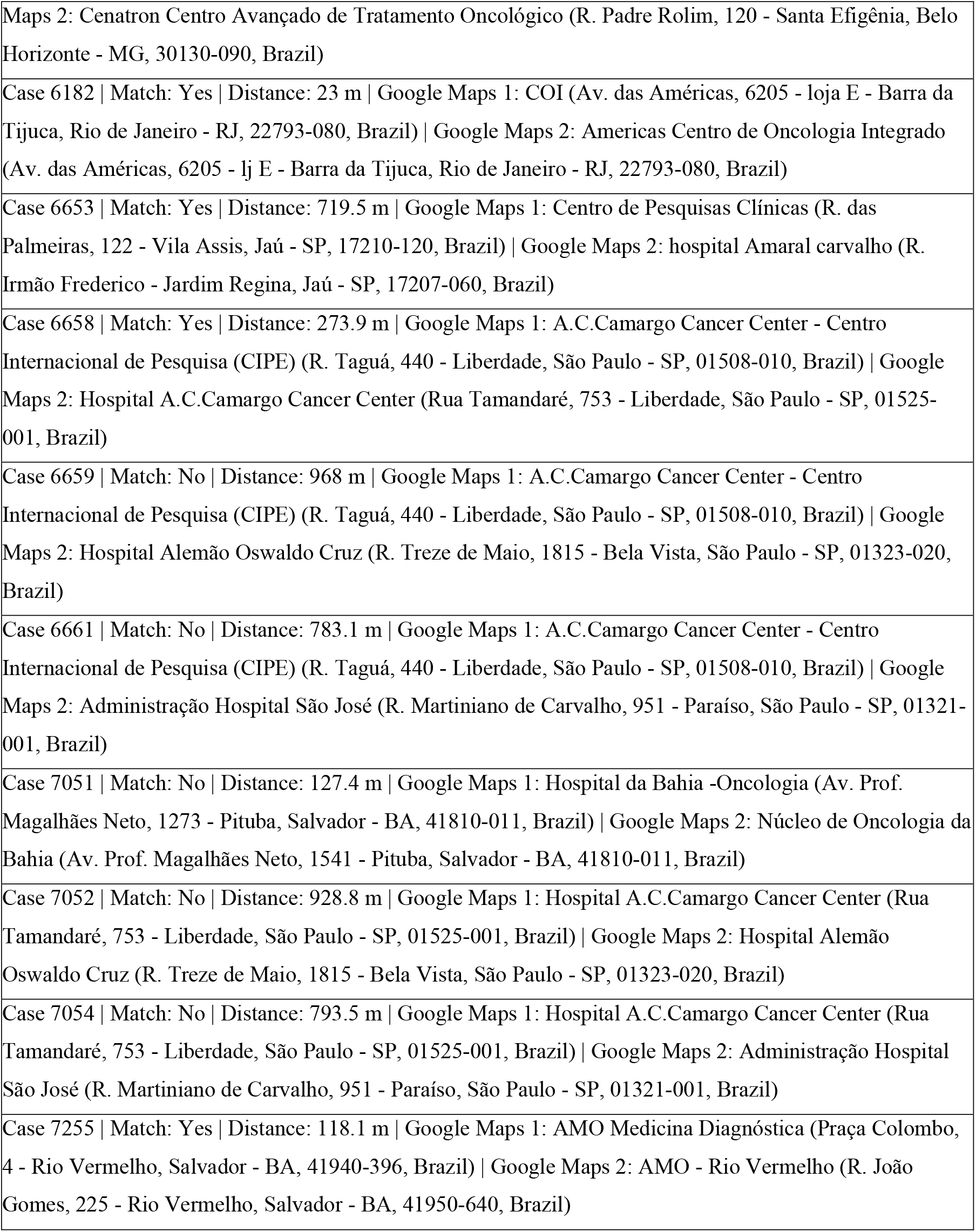
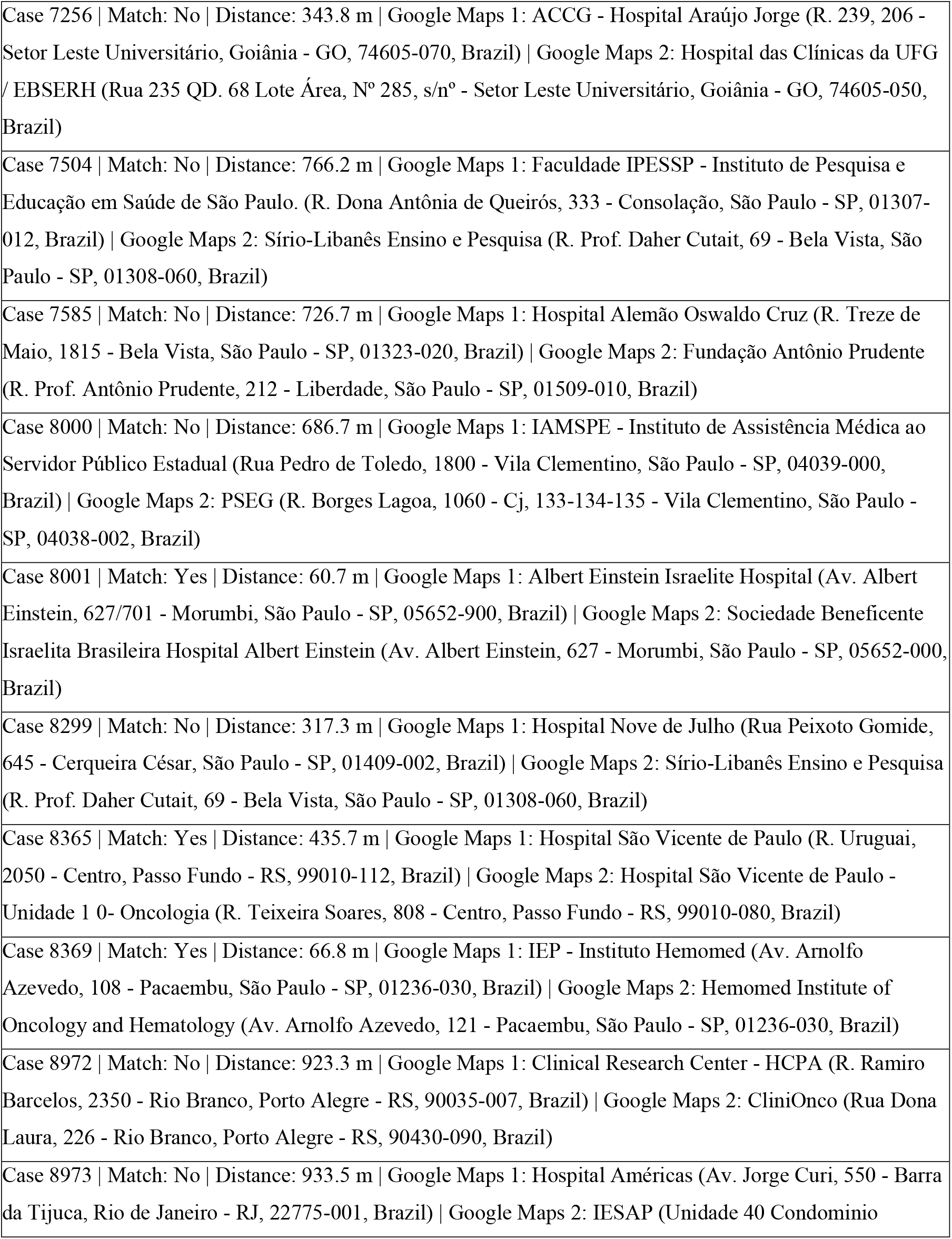
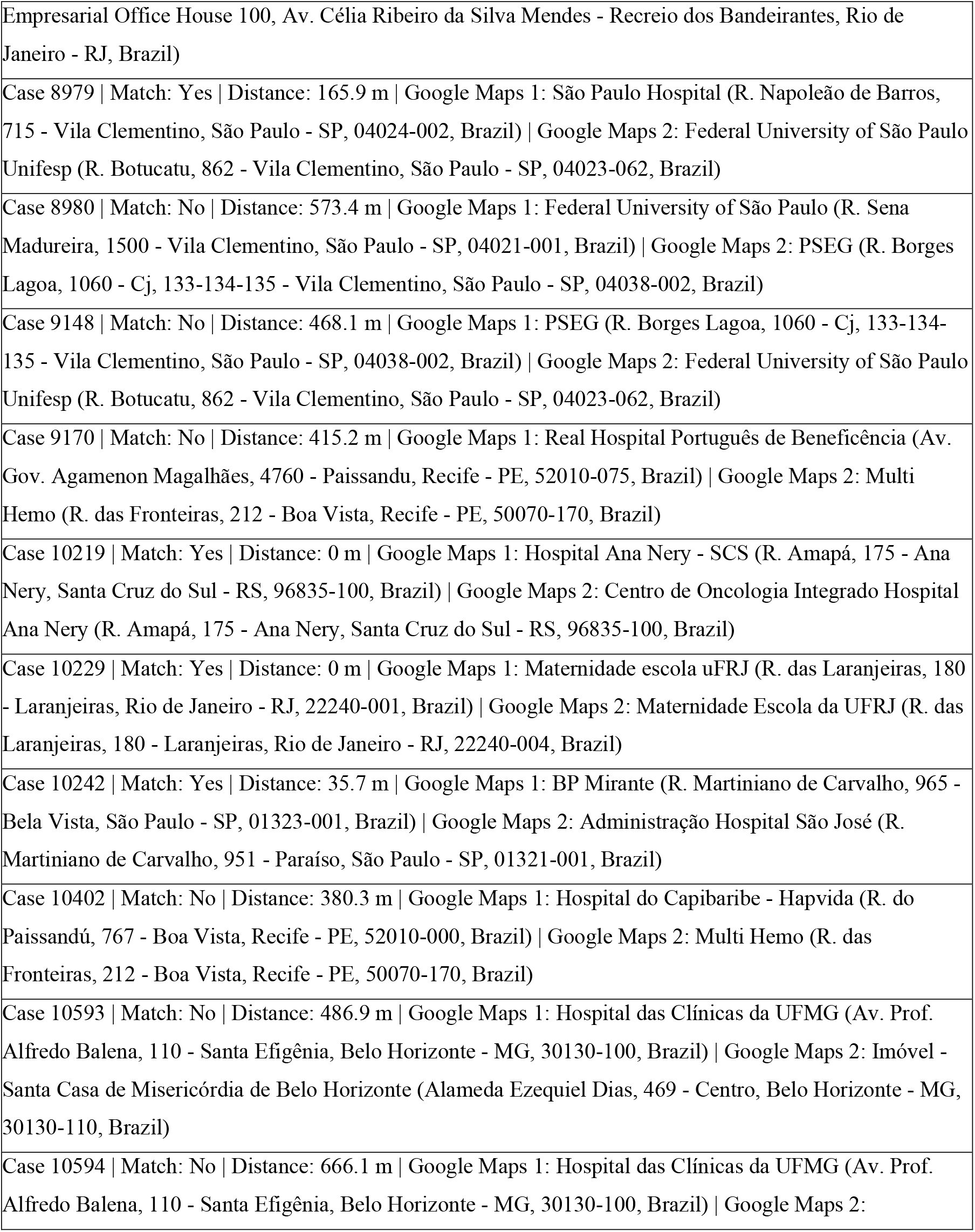
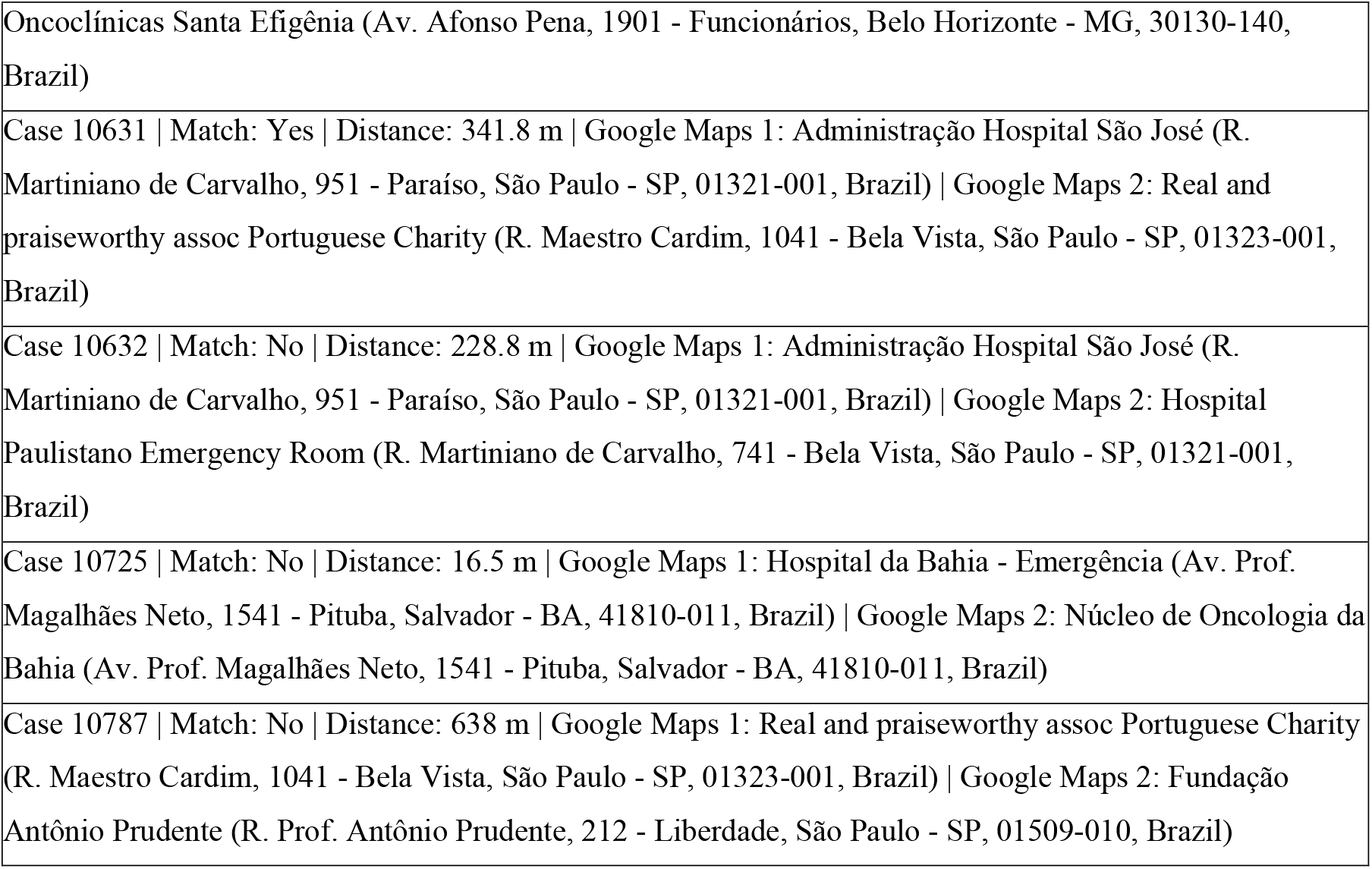
A random sample of 80 Google Maps Places pairs from Brazil, each located within 1,000 meters and possessing distinct Google Places Identifications, was classified by the authors with “Yes” or “No” match labels. This dataset served as the gold standard for prompt engineering and accuracy evaluation.

**SUPPL. TABLE 2.**
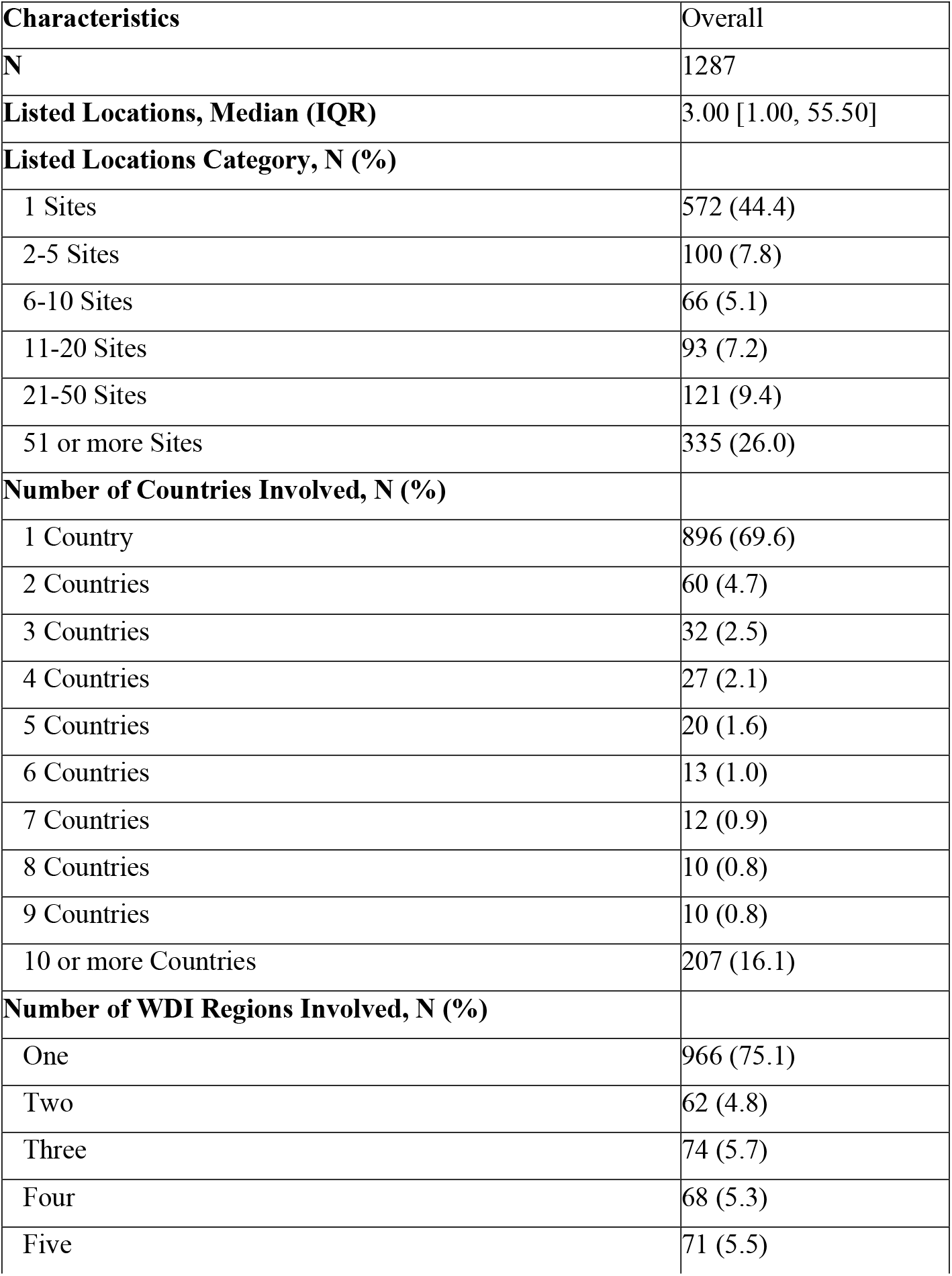
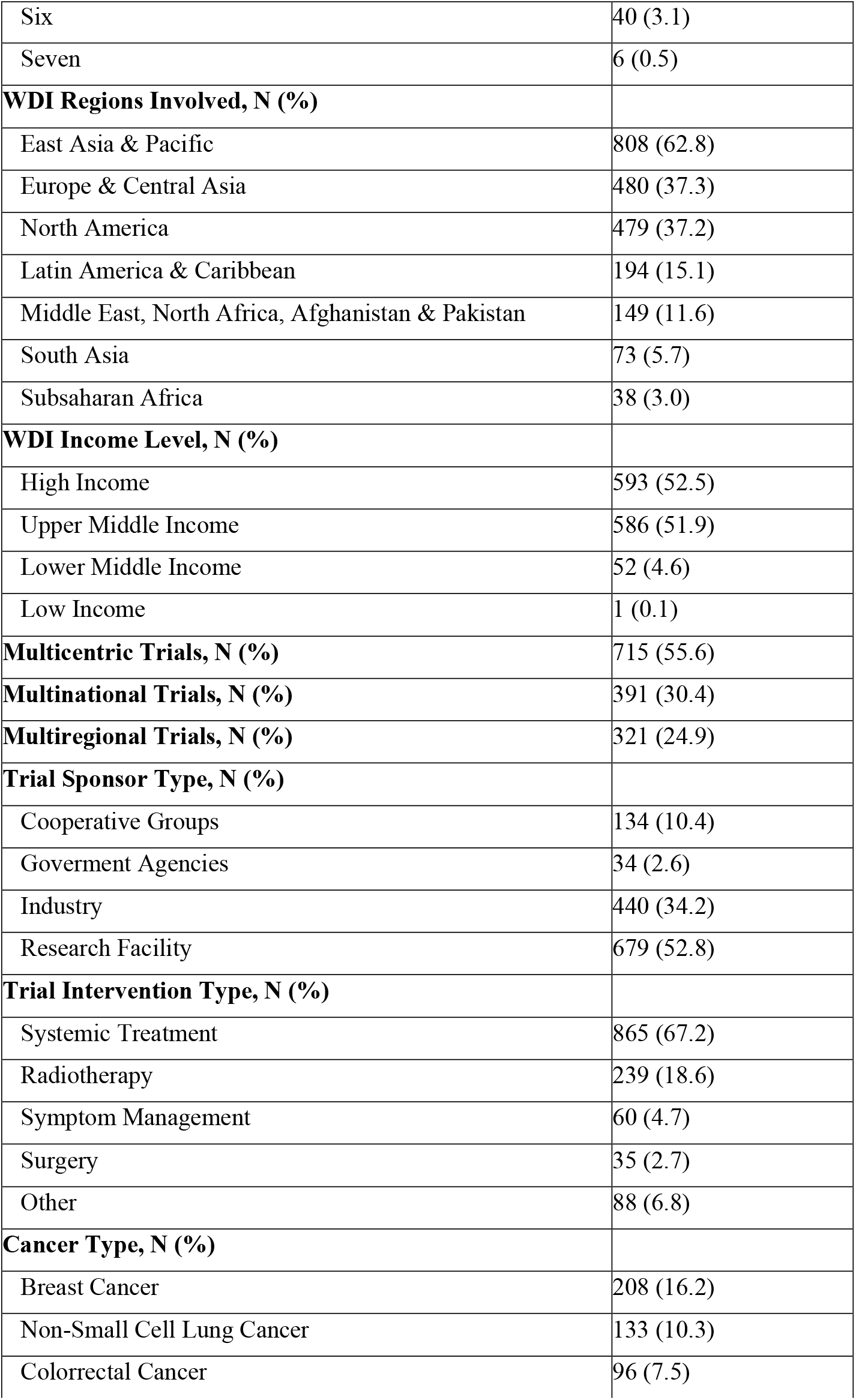
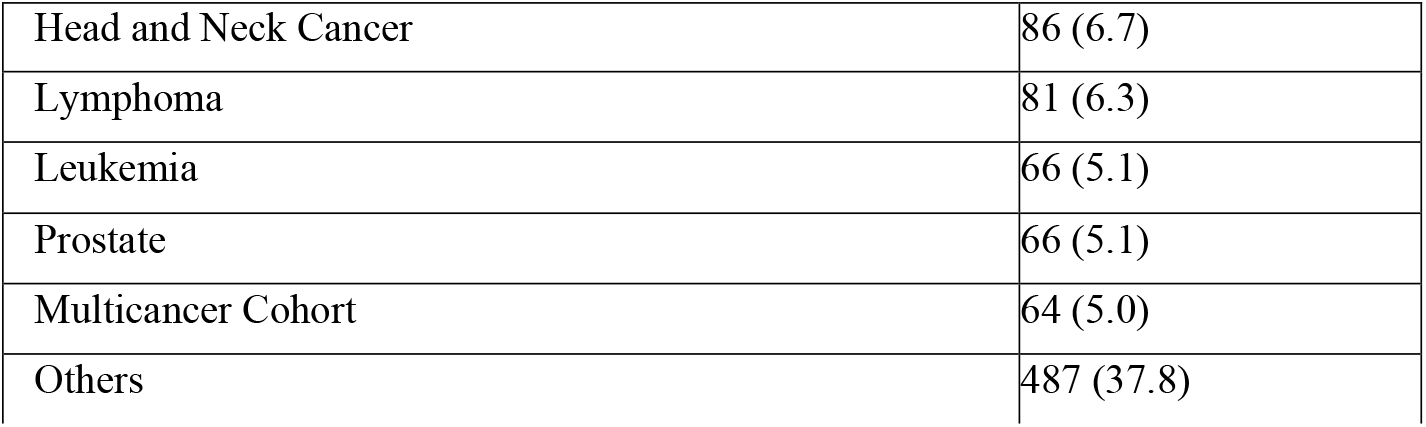
Characteristics of the Phase III Clinical Trials Included. Baseline characteristics of 1,287 Phase III clinical trials included in the analysis. Geographic distribution is presented by trial sites, countries, and World Bank Development Indicators (WDI) regions and income levels. Continuous variables are presented as median (interquartile range); categorical variables as N (%).

| **Characteristics** | **Overall** |
| --- | --- |
| **N** | 1287 |
| **Listed Locations, Median (IQR)** | 3.00 [1.00, 55.50] |
| **Listed Locations Category, N (%)** |  |
| 1 Sites | 572 (44.4) |
| 2-5 Sites | 100 (7.8) |
| 6-10 Sites | 66 (5.1) |
| 11-20 Sites | 93 (7.2) |
| 21-50 Sites | 121 (9.4) |
| 51 or more Sites | 335 (26.0) |
| **Number of Countries Involved, N (%)** |  |
| 1 Country | 896 (69.6) |
| 2 Countries | 60 (4.7) |
| 3 Countries | 32 (2.5) |
| 4 Countries | 27 (2.1) |
| 5 Countries | 20 (1.6) |
| 6 Countries | 13 (1.0) |
| 7 Countries | 12 (0.9) |
| 8 Countries | 10 (0.8) |
| 9 Countries | 10 (0.8) |
| 10 or more Countries | 207 (16.1) |
| **Number of WDI Regions Involved, N (%)** |  |
| One | 966 (75.1) |
| Two | 62 (4.8) |
| Three | 74 (5.7) |
| Four | 68 (5.3) |
| Five | 71 (5.5) |
| Six | 40 (3.1) |
| Seven | 6 (0.5) |
| **WDI Regions Involved, N (%)** |  |
| East Asia & Pacific | 808 (62.8) |
| Europe & Central Asia | 480 (37.3) |
| North America | 479 (37.2) |
| Latin America & Caribbean | 194 (15.1) |
| Middle East, North Africa, Afghanistan & Pakistan | 149 (11.6) |
| South Asia | 73 (5.7) |
| Subsaharan Africa | 38 (3.0) |
| **WDI Income Level, N (%)** |  |
| High Income | 593 (52.5) |
| Upper Middle Income | 586 (51.9) |
| Lower Middle Income | 52 (4.6) |
| Low Income | 1 (0.1) |
| **Multicentric Trials, N (%)** | 715 (55.6) |
| **Multinational Trials, N (%)** | 391 (30.4) |
| **Multiregional Trials, N (%)** | 321 (24.9) |
| **Trial Sponsor Type, N (%)** |  |
| Cooperative Groups | 134 (10.4) |
| Government Agencies | 34 (2.6) |
| Industry | 440 (34.2) |
| Research Facility | 679 (52.8) |
| **Trial Intervention Type, N (%)** |  |
| Systemic Treatment | 865 (67.2) |
| Radiotherapy | 239 (18.6) |
| Symptom Management | 60 (4.7) |
| Surgery | 35 (2.7) |
| Other | 88 (6.8) |
| **Cancer Type, N (%)** |  |
| Breast Cancer | 208 (16.2) |
| Non-Small Cell Lung Cancer | 133 (10.3) |
| Colorrectal Cancer | 96 (7.5) |
| Head and Neck Cancer | 86 (6.7) |
| Lymphoma | 81 (6.3) |
| Leukemia | 66 (5.1) |
| Prostate | 66 (5.1) |
| Multicancer Cohort | 64 (5.0) |
| Others | 487 (37.8) |

**SUPPL. TABLE 3.** Geographic distribution of 572 single-center Phase III clinical trials by World Bank Development Indicators (WDI) region, income level, and country. Categorical variables are presented as N (%).

| Characteristics | Overall |
| --- | --- |
| **N** | 572 |
| **WDI Regions Involved, N (%)** |  |
| East Asia & Pacific | 398 (69.6) |
| Europe & Central Asia | 73 (12.8) |
| Latin America & Caribbean | 8 (1.4) |
| Middle East, North Africa, Afghanistan & Pakistan | 20 (3.5) |
| North America | 59 (10.3) |
| South Asia | 14 (2.4) |
| **WDI Income Level, N (%)** |  |
| High income | 151 (26.5) |
| Upper middle income | 388 (68.2) |
| Lower middle income | 29 (5.1) |
| Low income | 1 (0.2) |
| **Country, N (%)** |  |
| China | 375 (65.6) |
| United States | 50 (8.7) |
| France | 19 (3.3) |
| Egypt | 15 (2.6) |
| India | 14 (2.4) |
| Korea, Republic of | 13 (2.3) |
| Canada | 9 (1.6) |
| Russian Federation | 9 (1.6) |
| Germany | 8 (1.4) |
| Italy | 8 (1.4) |
| United Kingdom | 5 (0.9) |
| Mexico | 4 (0.7) |
| Netherlands | 4 (0.7) |
| Brazil | 3 (0.5) |
| Japan | 3 (0.5) |
| Sweden | 3 (0.5) |
| Taiwan | 3 (0.5) |
| Czechia | 2 (0.3) |
| Denmark | 2 (0.3) |
| Hong Kong | 2 (0.3) |
| Norway | 2 (0.3) |
| Spain | 2 (0.3) |
| Argentina | 1 (0.2) |
| Armenia | 1 (0.2) |
| Austria | 1 (0.2) |
| Belgium | 1 (0.2) |
| Finland | 1 (0.2) |
| Georgia | 1 (0.2) |
| Hungary | 1 (0.2) |
| Iran, Islamic Republic of | 1 (0.2) |
| Ireland | 1 (0.2) |
| Israel | 1 (0.2) |
| Kuwait | 1 (0.2) |
| Malaysia | 1 (0.2) |
| Saudi Arabia | 1 (0.2) |
| Singapore | 1 (0.2) |
| Switzerland | 1 (0.2) |
| Syrian Arab Republic | 1 (0.2) |
| Ukraine | 1 (0.2) |

**SUPPL. TABLE 4.** Geographic distribution of 391 multinational Phase III clinical trials by World Bank Development Indicators (WDI) region and income level. Categorical variables are presented as N (%).

| **Characteristics** | **Overall** |
| --- | --- |
| **N** | 391 |
| **WDI Regions Involved, N (%)** |  |
| Latin America & Caribbean | 184 (47.1) |
| North America | 317 (81.1) |
| Europe & Central Asia | 316 (80.8) |
| East Asia & Pacific | 282 (72.1) |
| Subsaharan Africa | 38 (9.7) |
| South Asia | 59 (15.1) |
| Middle East, North Africa, Afghanistan & Pakistan | 129 (33.0) |
| **WDI Income Level, N (%)** |  |
| High income | 238 (99.6) |
| Upper middle income | 80 (33.5) |
| Lower middle income | 23 (9.6) |
| Low income | 239 (100.0) |

**SUPPL. TABLE 5.**
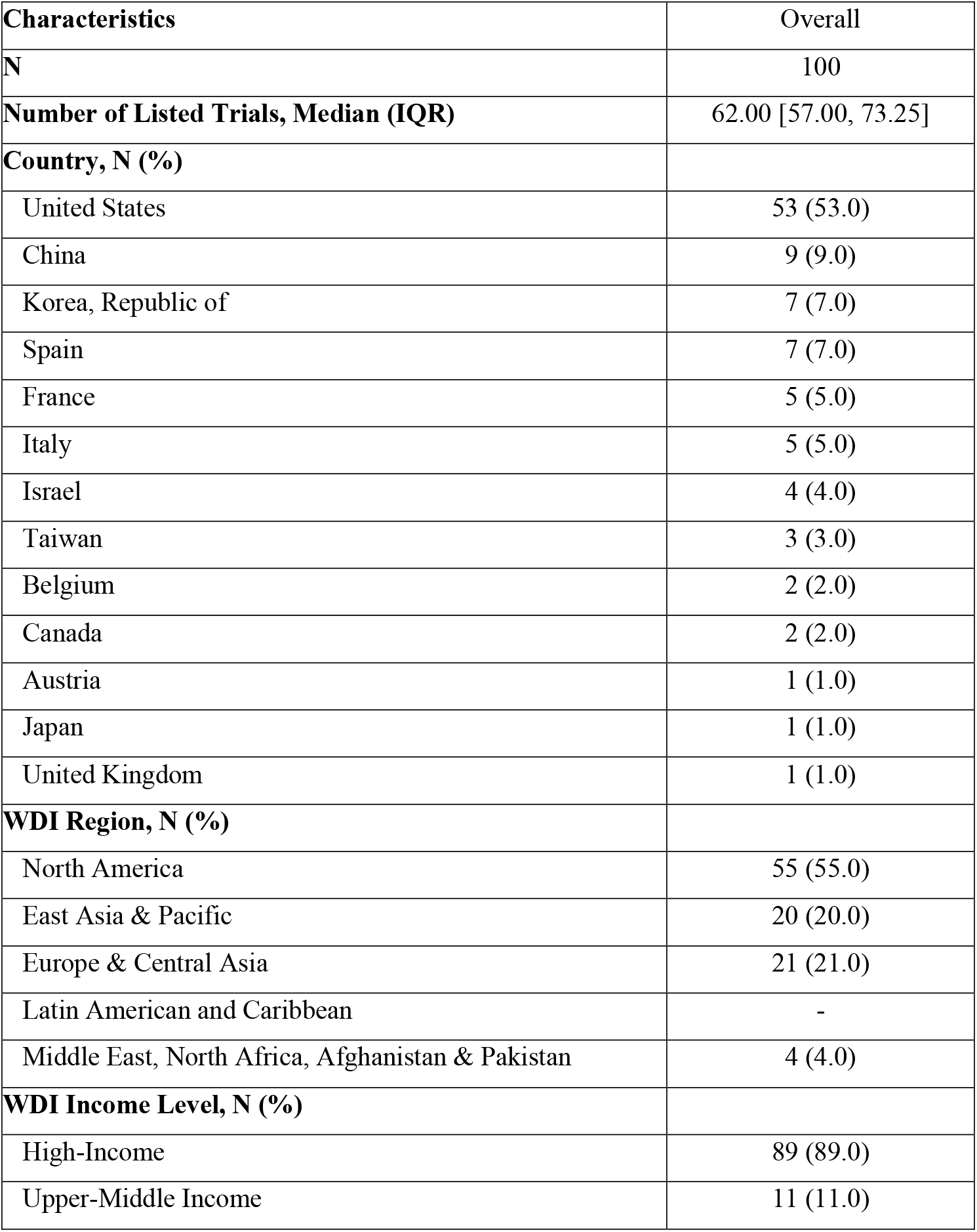
Characteristics of the 100 research facilities with the highest number of Phase III clinical trial participations, stratified by country, World Bank Development Indicators (WDI) region, and income level. Continuous variables are presented as median [interquartile range]; categorical variables as N (%).

**SUPPL. TABLE 6.** Characteristics of the 100 research facilities with the highest number of Phase III clinical trial participations, stratified by country, World Bank Development Indicators (WDI) region, and income level in the main and sensitivity analysis. Continuous variables are presented as median [interquartile range]; categorical variables as N (%).

| **Characteristics** | **Main Analysis** | **Sensitivity Analysis** |
| --- | --- | --- |
| **N** | 100 | 100 |
| **Number of Listed Trials, Median (IQR)** | 62.00 [57.00, 73.25] | 70.00 [62.75, 81.50] |
| **Country, N (%)** |  |  |
| United States | 53 (53.0) | 43 (43.0) |
| China | 9 (9.0) | 11 (11.0) |
| Korea, Republic of | 7 (7.0) | 7 (7.0) |
| Spain | 7 (7.0) | 8 (8.0) |
| France | 5 (5.0) | 6 (6.0) |
| Italy | 5 (5.0) | 5 (5.0) |
| Israel | 4 (4.0) | 5 (5.0) |
| Taiwan | 3 (3.0) | 4 (4.0) |
| Belgium | 2 (2.0) | 2 (2.0) |
| Canada | 2 (2.0) | 3 (3.0) |
| Austria | 1 (1.0) | - |
| Japan | 1 (1.0) | 3 (3.0) |
| United Kingdom | 1 (1.0) | 2 (2.0) |
| Germany | - | 1 (1.0) |
| **WDI Region, N (%)** |  |  |
| North America | 55 (55.0) | 46 (46.0) |
| East Asia & Pacific | 20 (20.0) | 25 (25.0) |
| Europe & Central Asia | 21 (21.0) | 24 (24.0) |
| Latin American and Caribbean | - | - |
| Middle East, North Africa, Afghanistan & Pakistan | 4 (4.0) | 5 (5.0) |
| **WDI Income Level, N (%)** |  |  |
| High-Income | 91 (91.0) | 89 (89.0) |
| Upper-Middle Income | 9 (9.0) | 11 (11.0) |

**SUPPL. TABLE 7.**
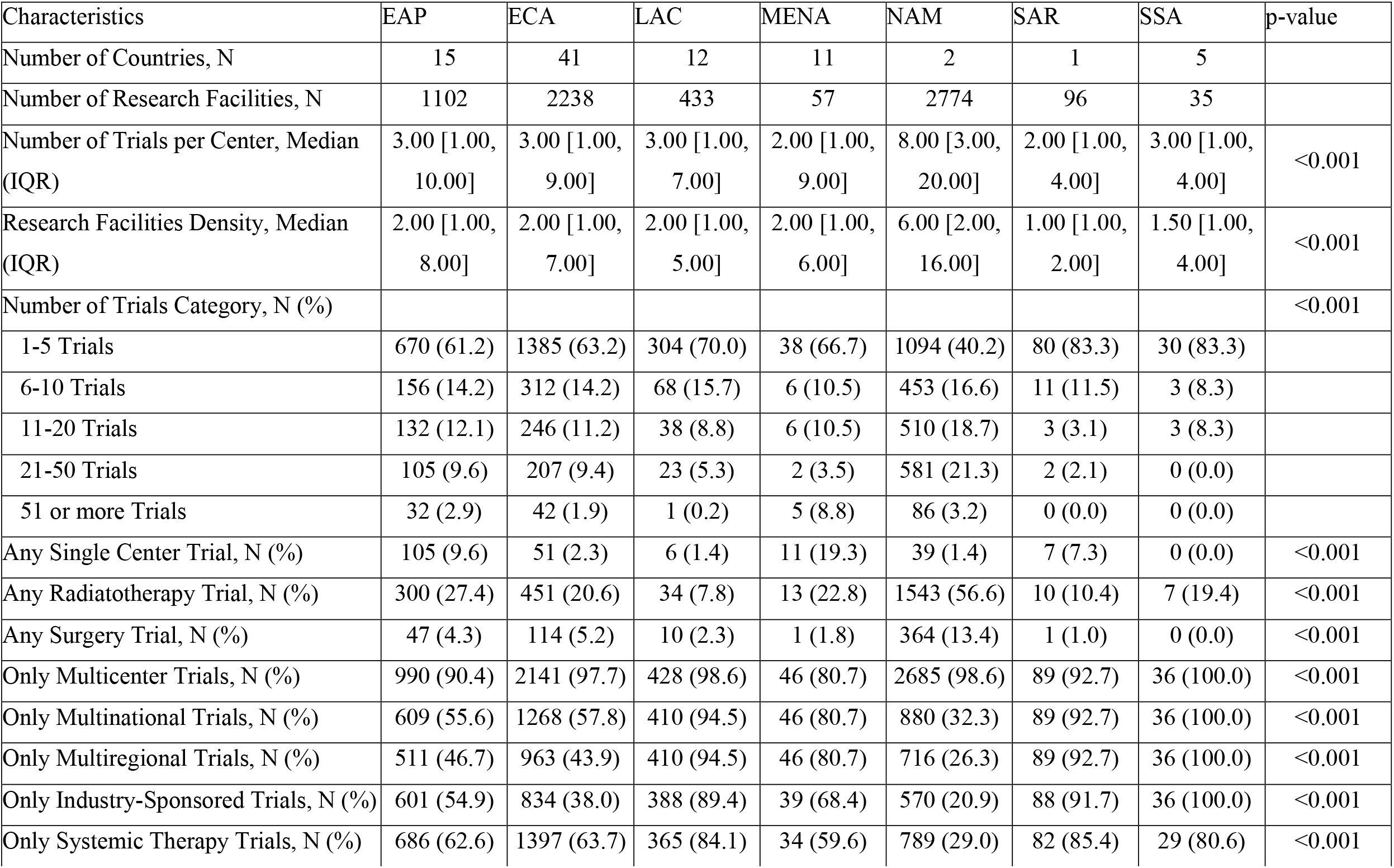
Comparison of Research Facility Characteristics by World Bank Development Indicators Region in the Sensitivity Analysis. P-values are from Kruskal-Wallis test for continuous variables and Fisher exact test for categorical variables. EAP = East Asia & Pacific; ECA = Europe & Central Asia; LAC = Latin America & Caribbean; MENA = Middle East, North Africa, Afghanistan & Pakistan; NAM = North America; SAR = South Asia; SSA = Sub-Saharan Africa.

| Characteristics | EAP | ECA | LAC | MENA | NAM | SAR | SSA | p-value |
| --- | --- | --- | --- | --- | --- | --- | --- | --- |
| Number of Countries, N | 15 | 41 | 12 | 11 | 2 | 1 | 5 |  |
| Number of Research Facilities, N | 1102 | 2238 | 433 | 57 | 2774 | 96 | 35 |  |
| Number of Trials per Center, Median (IQR) | 3.00 [1.00, 10.00] | 3.00 [1.00, 9.00] | 3.00 [1.00, 7.00] | 2.00 [1.00, 9.00] | 8.00 [3.00, 20.00] | 2.00 [1.00, 4.00] | 3.00 [1.00, 4.00] | <0.001 |
| Research Facilities Density, Median (IQR) | 2.00 [1.00, 8.00] | 2.00 [1.00, 7.00] | 2.00 [1.00, 5.00] | 2.00 [1.00, 6.00] | 6.00 [2.00, 16.00] | 1.00 [1.00, 2.00] | 1.50 [1.00, 4.00] | <0.001 |
| Number of Trials Category, N (%) |  |  |  |  |  |  |  | <0.001 |
| 1-5 Trials | 670 (61.2) | 1385 (63.2) | 304 (70.0) | 38 (66.7) | 1094 (40.2) | 80 (83.3) | 30 (83.3) |  |
| 6-10 Trials | 156 (14.2) | 312 (14.2) | 68 (15.7) | 6 (10.5) | 453 (16.6) | 11 (11.5) | 3 (8.3) |  |
| 11-20 Trials | 132 (12.1) | 246 (11.2) | 38 (8.8) | 6 (10.5) | 510 (18.7) | 3 (3.1) | 3 (8.3) |  |
| 21-50 Trials | 105 (9.6) | 207 (9.4) | 23 (5.3) | 2 (3.5) | 581 (21.3) | 2 (2.1) | 0 (0.0) |  |
| 51 or more Trials | 32 (2.9) | 42 (1.9) | 1 (0.2) | 5 (8.8) | 86 (3.2) | 0 (0.0) | 0 (0.0) |  |
| Any Single Center Trial, N (%) | 105 (9.6) | 51 (2.3) | 6 (1.4) | 11 (19.3) | 39 (1.4) | 7 (7.3) | 0 (0.0) | <0.001 |
| Any Radiotherapy Trial, N (%) | 300 (27.4) | 451 (20.6) | 34 (7.8) | 13 (22.8) | 1543 (56.6) | 10 (10.4) | 7 (19.4) | <0.001 |
| Any Surgery Trial, N (%) | 47 (4.3) | 114 (5.2) | 10 (2.3) | 1 (1.8) | 364 (13.4) | 1 (1.0) | 0 (0.0) | <0.001 |
| Only Multicenter Trials, N (%) | 990 (90.4) | 2141 (97.7) | 428 (98.6) | 46 (80.7) | 2685 (98.6) | 89 (92.7) | 36 (100.0) | <0.001 |
| Only Multinational Trials, N (%) | 609 (55.6) | 1268 (57.8) | 410 (94.5) | 46 (80.7) | 880 (32.3) | 89 (92.7) | 36 (100.0) | <0.001 |
| Only Multiregional Trials, N (%) | 511 (46.7) | 963 (43.9) | 410 (94.5) | 46 (80.7) | 716 (26.3) | 89 (92.7) | 36 (100.0) | <0.001 |
| Only Industry-Sponsored Trials, N (%) | 601 (54.9) | 834 (38.0) | 388 (89.4) | 39 (68.4) | 570 (20.9) | 88 (91.7) | 36 (100.0) | <0.001 |
| Only Systemic Therapy Trials, N (%) | 686 (62.6) | 1397 (63.7) | 365 (84.1) | 34 (59.6) | 789 (29.0) | 82 (85.4) | 29 (80.6) | <0.001 |

**SUPPL. TABLE 8.** Multilevel Logistic Regression Analysis of Predictors of Unidentified Trial Site Locations. Results from univariable and multivariable multilevel logistic regression models examining predictors of unidentified trial site locations for main analysis and sensitivity analysis. Odds ratios (OR) and 95% confidence intervals (95% CI) are presented. Reference categories: North America (NAM) for WDI region; government-sponsored trials for sponsor type.

| Analysis | Variable | Univariable OR | Multivariable OR |
| --- | --- | --- | --- |
| **Main Analysis** | **WDI Region** |  |  |
|  | North America | Ref. | Ref. |
|  | East Asia & Pacific | 0.57 (95%CI 0.48–0.66) | 0.56 (95%CI 0.48–0.66) |
|  | Europe & Central Asia | 2.08 (95%CI 1.84–2.36) | 2.07 (95%CI 1.83–2.34) |
|  | Latin America & Caribbean | 4.46 (95%CI 3.82–5.21) | 4.41 (95%CI 3.77–5.14) |
|  | Middle East, North Africa, Afghanistan & Pakistan | 0.37 (95%CI 0.23–0.59) | 0.38 (95%CI 0.24–0.60) |
|  | South Asia | 2.37 (95%CI 1.60–3.53) | 2.39 (95%CI 1.61–3.55) |
|  | Subsaharan Africa | 0.75 (95%CI 0.39–1.45) | 0.76 (95%CI 0.39–1.46) |
|  | **Sponsor** |  |  |
|  | Government Agencies | Ref. | Ref. |
|  | Cooperative Groups | 1.03 (95%CI 0.32–3.27) | 1.02 (95%CI 0.25–4.23) |
|  | Industry | 15.22 (95%CI 5.35–43.35) | 17.13 (95%CI 4.64–63.24) |
|  | Research Facility | 1.66 (95%CI 0.57–4.85) | 2.09 (95%CI 0.56–7.78) |
| **Sensitivity Analysis** | **WDI Region** |  |  |
|  | North America | Ref. | Ref. |
|  | East Asia & Pacific | 0.74 (95%CI 0.67–0.82) | 0.73 (95%CI 0.66–0.81) |
|  | Europe & Central Asia | 0.92 (95%CI 0.84–1.01) | 0.91 (95%CI 0.83–0.99) |
|  | Latin America & Caribbean | 1.54 (95%CI 1.36–1.76) | 1.52 (95%CI 1.34–1.72) |
|  | Middle East, North Africa, Afghanistan & Pakistan | 0.41 (95%CI 0.29–0.58) | 0.42 (95%CI 0.30–0.59) |
|  | South Asia | 1.08 (95%CI 0.85–1.38) | 1.08 (95%CI 0.85–1.38) |
|  | Subsaharan Africa | 1.34 (95%CI 0.86–2.10) | 1.36 (95%CI 0.87–2.12) |
|  | **Sponsor** |  |  |
|  | Government Agencies | Ref. | Ref. |
|  | Cooperative Groups | 1.44 (95%CI 0.70–2.97) | 1.52 (95%CI 0.65–3.58) |
|  | Industry | 9.06 (95%CI 4.62–17.79) | 10.39 (95%CI 4.74–22.76) |
|  | Research Facility | 3.24 (95%CI 1.60–6.57) | 3.95 (95%CI 1.76–8.90) |

